# The Genetics of Fibromyalgia and its Relationships to Psychiatric and Medical Traits

**DOI:** 10.1101/2025.09.18.25335991

**Authors:** Uri Bright, Sarah Beck, Daniel F. Levey, Joseph D. Deak, The VA Million Veteran Program, J. Michael Gaziano, Murray B. Stein, Joel Gelernter

## Abstract

Fibromyalgia is a chronic heritable syndrome, with little prior genetic work taking a syndromic approach. In this study we aimed to discover genetic mechanisms underlying fibromyalgia. We conducted genome-wide association studies (GWAS) of fibromyalgia in subjects of European (EUR), African (AFR) and Latin American (AMR) ancestries, combining data from several samples (total N, 85,139 cases; 1,642,433 controls). We also conducted a multi-trait analysis of GWAS (MTAG), leveraging pain summary statistics to achieve enhanced power for fibromyalgia analyses. We then applied a series of post-GWAS methods to analyze the genetic association between fibromyalgia and a variety of psychological and physiological phenotypes. We found 10 genome-wide significant (GWS) loci associated with fibromyalgia in EUR, 1 in AFR, 12 cross-ancestry, and 45 in the EUR MTAG. Most of these loci were previously associated with pain, cognitive function, autoimmune response, or general health measures like BMI and blood pressure. Genetic correlation analysis revealed a moderate negative correlation with physical activity, and strong genetic correlations with chronic pain, PTSD and depression (r_g_ ≥ 0.69). Genomic structural equation modeling helped to place fibromyalgia in the context of a set of psychiatric, medical, and lifestyle traits. Additional findings regarding local genetic correlations and genetic causality point to genetic mechanisms that provide a strong basis for some of the main characteristics of fibromyalgia and its comorbidities. These findings provide potential targets for future studies to improve diagnosis and treatment of fibromyalgia.

## Background

Fibromyalgia is a chronic disorder characterized by widespread pain, muscle stiffness, sleep disturbances, fatigue, depression, and anxiety [1]. The point prevalence of fibromyalgia in the world population is 2.7%, with females making up 75% of cases [2]. There are no pathognomonic tests (e.g., blood-based or imaging biomarkers), and its diagnosis is thus based on identifying symptoms that are associated with the disease and exclusion of alternative explanations [1].

Fibromyalgia is influenced by genetic risk factors [3]: the risk of fibromyalgia among relatives of subjects who suffer from the disorder is eight times higher than in the general population [4], and it may reach to 13-fold in siblings [5]. Genome-wide association studies (GWAS) have revealed dozens of genes associated with chronic pain and other pain-related disorders [6]. Two fibromyalgia-specific GWAS have been published to date; both used relatively small sample sizes and found no significant loci in European (EUR) or African (AFR) populations [7, 8]. A study that examined the genetic basis of fibromyalgia as part of a long list of pain disorders did also not find any significant variant specifically related to fibromyalgia [9]. A GWAS of chronic widespread musculoskeletal pain, a symptom of fibromyalgia, reported three genome-wide associations in *RNF123*, *ATP2C1* and *COMT* [10].

A phenome-wide association study (pheWAS) revealed significant associations between fibromyalgia and 304 phenotypes (out of 1,740 phecodes tested); the strongest of these associations were with pain phenotypes (e.g. chronic pain, joint pain), traits directly related to pain (e.g. osteoarthrosis, spondylosis, migraine), autoimmune disease (erythematosus lupus) and psychiatric disorders (e.g. depression) [8]. Indeed, there is a strong association between fibromyalgia and psychiatric disorders: the lifetime prevalences of depression, anxiety, posttraumatic stress disorder (PTSD) and attention-deficit/hyperactivity disorder (ADHD) among fibromyalgia patients are reported to be 74%, 60%, 57% and 45%, respectively [11–13].

The therapeutic approach typically includes a combination of psychotherapy, pharmacology (sometimes off-label; there are also three FDA-approved medications) and lifestyle changes [2]. The latter often involves the introduction or increased frequency of aerobic physical exercise, a first-line treatment for the disorder. The pharmacological treatments for fibromyalgia tend to be based on antidepressant and muscle-relaxant drugs, while the efficacy of opioids is quite limited [2, 14]. Several small studies suggest that cannabis may be helpful to alleviate pain in fibromyalgia patients [15]. These treatments are variably effective; new therapeutic approaches are needed.

Currently, it is unclear whether fibromyalgia can be prevented, and the common treatments are often unsatisfactory [16]. In this study, we aimed to discover genetic mechanisms that may identify novel treatment targets and open new paths for research regarding possible ways to alleviate symptoms of this disorder, that impairs the quality of life of so many people worldwide. Further, we hope to contribute to better prediction of who is at risk for the development of fibromyalgia which could lead to discoveries about prevention. To that cause, we conducted a meta-analyzed GWAS in individuals of EUR, AFR and Latin American (AMR) ancestries, as well as a cross-ancestry and sex-stratified analyses, based on several different large samples including All of US (AoU) [17], the Million Veteran Program (MVP) [18], the UK Biobank (UKBB) [19] and Finngen [20] biobanks. We also conducted post-GWAS analyses to assess gene-based associations, transcriptome-wide associations, global and local genetic correlations, and genetic causality between fibromyalgia and a list of autoimmune diseases, psychiatric traits and general health phenotypes that are assumed to be associated with fibromyalgia [1, 2, 21]. Finally, we conducted genomic structural equation modeling (gSEM), to understand the genomic relationships between fibromyalgia and other psychiatric, autoimmune, general health and pain-related traits.

## Methods

### Cohorts

We analyzed data from All of Us, version 8 (AoU), the Million Veteran Program (MVP), and the UK Biobank (UKBB). Genotyping and quality control procedures of AoU, MVP and UKBB (EUR1 and EUR2) were previously described in refs. [22], [18], [23] and [24], respectively.

### Initial Analyses for Phenotype Definition

To include only confirmed cases of a studied disorder, a phenotype is often defined by a stringent ICD-10 code-based phenotype definition (a): ≥ 2 outpatient and/or 1 inpatient visit/s. In this study, to create well-powered analyses, we aimed to assemble a larger sample size by considering inclusion of (b) fibromyalgia subjects meeting a less-stringent ICD-10 phenotype definition (≥ 1 outpatient visit and/or 1 inpatient visits), or (c) self-report (of having been diagnosed with fibromyalgia). To do this, we conducted separate GWAS of fibromyalgia, assigning subjects as cases according to the three different phenotype definitions: stringent ICD-10, less-stringent ICD-10 (with a single outpatient diagnosis considered sufficient to define affection), and self-report. We then calculated the genetic correlations between GWAS based on these differing case definitions, to ascertain genetic similarity between these phenotypes. We conducted these preliminary phenotype definition tests in each of AoU, MVP and UKBB separately.

In each of the cohorts, we defined fibromyalgia two or three ways, depending on data availability: a stringent ICD-10 code definition, less-stringent ICD-10 code definition and self-report. Subjects that fit more than one of these phenotypes were excluded from this part of the analysis completely, i.e. they were not defined as cases nor controls (subjects that fit, for example, both strict ICD-10 code and self-report, were excluded; this exclusion applied only for these analyses conducted to evaluate the genetic relationship of subjects defined by these three phenotype methods). Only subjects that fit one category but not the others were included in the analysis as cases of the relevant category. Subjects that did not fit any category were classified as controls. Any subject classified as a case in one of these analyses, was excluded from the others (i.e., there was no situation where a subject was used as a case for one analysis and as a control subject in another). We conducted GWAS of these phenotypes separately, removing one individual from each pair of related subjects (kinship coefficient cutoff = 0.1), retaining as many cases as possible. GWAS were conducted with PLINK 2.0 using logistic regression, with sex, age, and the first ten genetic principal components (PCs) as covariates:

In AoU, we extracted three different fibromyalgia phenotypes: stringent ICD-10 (n_cases_=2,932, n_controls_=212,390), less-stringent ICD-10 (n_cases_=1,951, n_controls_=210,571) and self-report (“Including yourself, who in your family has had fibromyalgia? Select all that apply. – Self”; n_cases_=5,339, n_controls_=208,813). In MVP, we extracted two different fibromyalgia phenotypes: stringent ICD-10 (n_cases_=24,041, n_controls_=384,196) and less-stringent ICD-10 (n_cases_=41,847, n_controls_=379,286). In UKBB, we extracted two different fibromyalgia phenotypes: stringent ICD-10 (EUR1: n_cases_=2,328, n_controls_=374,731; EUR2: n_cases_=329, n_controls_=52,072) and self-report (“Have you ever been told by a doctor that you have had any of the following conditions? - Fibromyalgia syndrome -Yes”; EUR1: n_cases_=1,666, n_controls_=374,840; EUR2: n_cases_=228, n_controls_=52,081), and conducted an inverse variance weighing meta-analysis of EUR1 and EUR2 results of each phenotype (using METAL [25]) before further analyses. For all the analyses we used LDSC [26] to calculate the liability-scaled heritability estimate (h^2^) and intercept (population prevalence was set at 2.7% [2]). We also tested the genetic correlations for the fibromyalgia phenotypes between these fibromyalgia case definitions, defined by stringent ICD-10 code definition, less-stringent ICD-10 code definition, and self-report. A strong genetic correlation between these sub-cohorts would confirm that these different definitions of fibromyalgia correspond to genetically similar traits, enabling the valid inclusion of subjects of these different cohorts. We also included in our analysis EUR fibromyalgia summary statistics from Finngen, release 12 [20], defined by the stringent ICD-10 code (n_cases_=3,623, n_controls_=357,579).

### Main Analysis

After we confirmed that the various fibromyalgia phenotypes (stringent ICD-10, less-stringent ICD-10 and self-report) are strongly genetically correlated (see previous section) – suggesting that they are likely representing a consistent trait – we defined a singular fibromyalgia phenotype in AoU, MVP and UKBB: subjects with stringent ICD-10, less-stringent ICD-10 and self-report definitions of fibromyalgia, including overlapping samples, were defined as cases. All other subjects were defined as controls. In AoU and MVP we conducted separate GWAS in subjects of European (EUR), African (AFR) and Latin American (AMR) genetically-defined ancestries. In UKBB we conducted the analysis in EUR. In each cohort, after removing one individual from each pair of related subjects (kinship coefficient cutoff = 0.1), retaining as many cases as possible, GWAS was conducted with PLINK 2.0 using logistic regression, with sex, age and the first ten genetic PCs as covariates. Variants with minor allele frequency (MAF) <0.1% and Hardy-Weinberg equilibrium (HWE) p<1×10^−6^ were excluded. In UKBB, we meta-analyzed EUR1 and EUR2 results before further analyses. For each ancestry in each cohort we also conducted sex-stratified GWAS. EUR, AFR, AMR and cross-ancestry SE-based meta-analyses (for all subjects and sex-stratified) were conducted using METAL [25]. The non-sex-stratified EUR meta-analysis included fibromyalgia summary statistics from the Finngen biobank [20]. In all the analyses we applied a standard genome-wide multiple testing correction (p<5×10^− 8^). The results were visualized in a Manhattan plot using the R package qqman [27]. Regional Manhattan plots were created using LocusZoom [28].

### FUMA and MAGMA gene-based and gene set analyses

We used FUMA [29] to define genomic risk loci and map the lead variants’ exact location in the genome. We used MAGMA [30], implemented in the FUMA platform, to conduct a gene-based analysis of the EUR, AFR, AMR and cross-ancestry meta-analyses. In the gene-based analyses, input SNPs were mapped to 19,069 protein-coding genes in EUR, 10,995 in AFR, 18,856 in AMR and 19,173 cross-ancestry, and corrected for false-discovery rate (FDR).

### Genetic Correlations and SNP-based heritability

We used linkage disequilibrium score regression (LDSC) [26] based on the linkage disequilibrium reference from the 1000 Genomes data [31] for all EUR cohorts. We calculated the liability-scaled SNP-based heritability estimate (h^2^) and intercept of fibromyalgia in every cohort separately (population prevalence was set on 2.7% [2]), and the genetic correlations (r_g_) between fibromyalgia phenotypes in the different cohorts. After meta-analysis, we calculated h^2^ for the meta-analyzed fibromyalgia trait and its r_g_ with a list of traits related to psychiatric disorders, auto immune diseases, sleep, and general health [20, 24, 32–45], phenotypically associated with fibromyalgia according to prior literature [1, 2]. We also conducted similar analyses for the sex-stratified meta-analyses of fibromyalgia in EUR and for the EUR MTAG, using the same summary statistics for the comparative traits that we used for the main analysis (we did not use sex-specific summary statistics of other traits due to low availability of well-powered GWAS of most of the traits we analyzed, and because we aimed to maintain the analyses with as few differences as possible, for better comparison between the results). After Bonferroni correction for 100 tests (4 fibromyalgia phenotypes x 25 traits), the statistical significance threshold was set at p=0.0005.

### Multi-Trait Analysis of GWAS (MTAG)

To enhance the statistical power of the EUR fibromyalgia GWAS we used MTAG [46], a method that enables joint analysis of traits that are genetically correlated with each other, and provides estimates of trait-specific effects. We used pain, a similar phenotype to the trait with the strongest genetic correlation with fibromyalgia we found using LDSC (chronic pain). A diagnosis of fibromyalgia may be categorized as pain, which raised the risk of using overlapping samples diagnosed under overlapping traits. Although MTAG is appropriate for overlapping samples, to minimize confounds we avoided using datasets with potentially high case overlap with fibromyalgia, and therefore we used summary statistics for pain among Finngen subjects, with 237,944 cases and 261,418 controls [20]. This pain phenotype was defined by 16 different diagnoses associated with limb, back, neck, head and abdominal pain. There were only 817 fibromyalgia subjects in the same group, <0.01% the number of the pain patients, illustrating the low susceptibility for case overlap. The r_g_ between these two traits was 0.72 (± 0.026). For an additional, more inclusive analysis, we aimed to capture the wider complexity of fibromyalgia as both a pain- and psychiatric-related trait. For this analysis we used depression for the MTAG analysis, because of the availability of a large-scaled GWAS of EUR MDD that does not include any of the cohorts we used in our study. We used MDD summary statistics from a large meta-analysis [47], taking a leave-one-out (LOO) approach, excluding subjects from MVP, UKBB, Finngen – and also 23andMe due to use restrictions. After exclusion, the MDD dataset included 181,654 MDD cases and 862,012 controls. The r_g_ between fibromyalgia and MDD was 0.63 (± 0.025), and between pain and MDD was 0.56 (± 0.021). In both sets of analysis, MTAG was conducted using fibromyalgia as the main trait, with variants restricted to only those common to the two GWAS, with MAF > 0.01. Because MDD had a relatively lower genetic correlation with fibromyalgia and with pain than the recommended threshold for MTAG (rg ≥ 0.7), we focused on the MTAG of fibromyalgia leveraged by pain (fibromyalgia MTAG leveraged by pain and MDD together is reported in the supplementary material).

### Mendelian Randomization (MR)

We conducted MR analyses to estimate the causality between fibromyalgia and 25 traits of interest (see previous section), assessing fibromyalgia both as exposure and outcome. All MR analyses were conducted using MRlap [48], which is appropriate for MR analysis with potentially overlapping cohorts. We ran the inverse variance weighted (IVW) model, with two p-value thresholdsDof 1×10^−5^ and 1×10^−8^ to select genetic instruments. Due to a low number of variants left after pruning using the stricter threshold of 1×10^−8^, we report only the 1×10^−5^ threshold analysis in our results (MR analysis using a p-value threshold of 1×10^−8^ is reported in the supplementary material). Instrument pruning was conducted based on an LD threshold of 0.05. For every analysis, MRlap performs a correction for overlapping samples and other potential biases such as outliers and presents the statistical difference between the observed and corrected values. Where the difference between these values was significant (p<0.05) we presented the corrected values in the results section; where the difference was not significant, we presented the observed values. Bonferroni correction for 100 tests yielded a statistical significance threshold of p=0.0005.

### Local Analysis of covariant Association (LAVA)

We used LAVA [49] to calculate local genetic correlations between fibromyalgia and 25 traits of interest in EUR. The genome was divided into 2,495 genomic regions, to provide minimal LD between the regions and maintain an approximately equal size of the regions of ∼1DMB. Breakpoints between regions were computed according to the LD between neighboring SNPs as described previously [49], maintain regions as relatively independent. Univariate local correlations were calculated for each trait, and only regions that reached significance (p=2 × 10^−5^ after Bonferroni correction) were used to calculate genetic correlations among the cohorts (8,384 regions in 25 pairs). Bonferroni correction for 8,384 tests yielded a statistical significance threshold of p=5.96 × 10^−6^.

### Transcriptome-Wide Association Study (TWAS)

We conducted TWAS using GTEx_v8 [50], which provides expression data of 49 tissues in EUR samples. We used the 1000 Genomes dataset as LD reference. Using FUSION [51], we identified associated genes, then processed the results to distinguish conditionally independent genes. In 49 tissues, there were a total of 300,187 genes measured (an average of 6,126 ± 2,787 per tissue). We therefore used a Bonferroni correction for 300,187 tests (0.05/300,187) to set a p-value threshold of 1.66×10^−7^.

### Genomic Structural Equation Modeling (gSEM)

We conducted a gSEM analysis [52] on fibromyalgia and 22 traits that had significant genetic correlations with fibromyalgia. Three of these traits – academic degree, executive functioning and physical activity, which had a negative genetic correlation with fibromyalgia – were reverse-coded before the analysis. Exploratory factor analysis (EFA) was performed followed by confirmatory factor analysis (CFA) to identify and confirm the factor structure. EFA models containing 1-9 factors were evaluated based upon eigenvalues, sum of squared (SS) loadings, cumulative variance explained, and the distribution of variance explained across the respective factors. CFA models were assessed using traditional fit indices [52].

### Drug Repurposing

We used the drug.MATADOR database, which is implemented in ShinyGo 0.82 [53], to identify potential drug-target interactions, using 173 genes that had a significant effect in at least one of the methods we used in this study – GWAS, MAGMA, MTAG and TWAS – in any of the ancestries we studied.

### Blood Type Phenotype

After we found a significant effect of *ABO* (a blood-type determining gene) in the MTAG, and based on known associations between blood type and pain perception [54, 55], we compared the ABO blood group blood type phenotype between fibromyalgia subjects and the general population, using data available in UKBB.

## Results

### Initial Analyses for Phenotype Definition

In each of the cohorts we analyzed in this study (AoU, MVP, UKBB), we conducted a separate GWAS for every phenotype definition of fibromyalgia (stringent ICD-10, less-stringent ICD-10, self-report), and then calculated the heritability and the genetic correlations between them (**Supplementary Table S1**). In AoU and MVP, we found significant genetic correlations between all the different phenotypes, with r_g_∼=1.00. In UKBB, the genetic correlation between the two defined phenotypes was not significant, likely due to relatively small sample sizes. We therefore tested the genetic correlation between the UKBB phenotypes and their closest equivalents in AoU (self-report in UKBB vs self-report in AoU, and stringent ICD-10 in UKBB vs stringent ICD-10 in AoU) and found a nominally significant effect of r_g_=0.77 between the self-report traits, and a significant effect for the stringent ICD-10 traits, with r_g_=1.07 (**Table S2**). In view of these results, we combined all the phenotypic variations and assembled inclusive fibromyalgia phenotypes in all cohorts (**Table 1**).

**Table 1:**
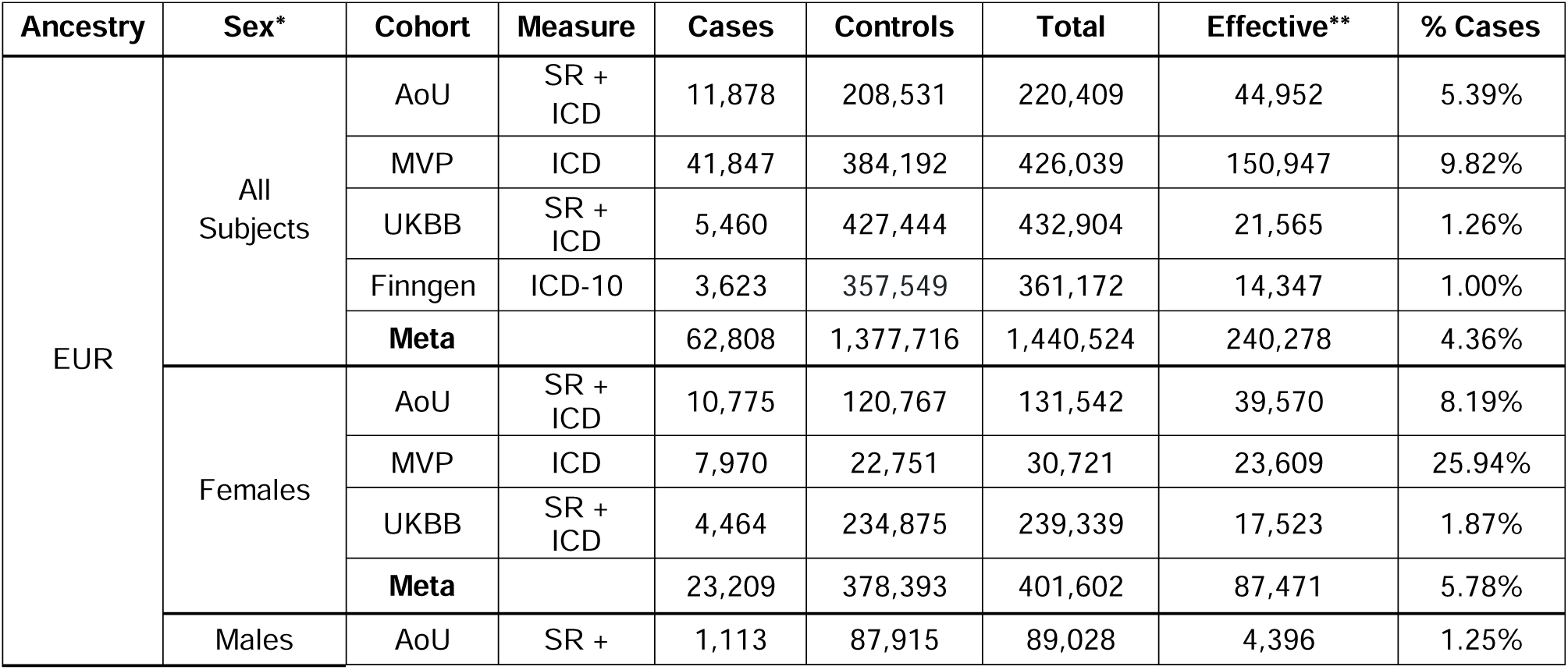
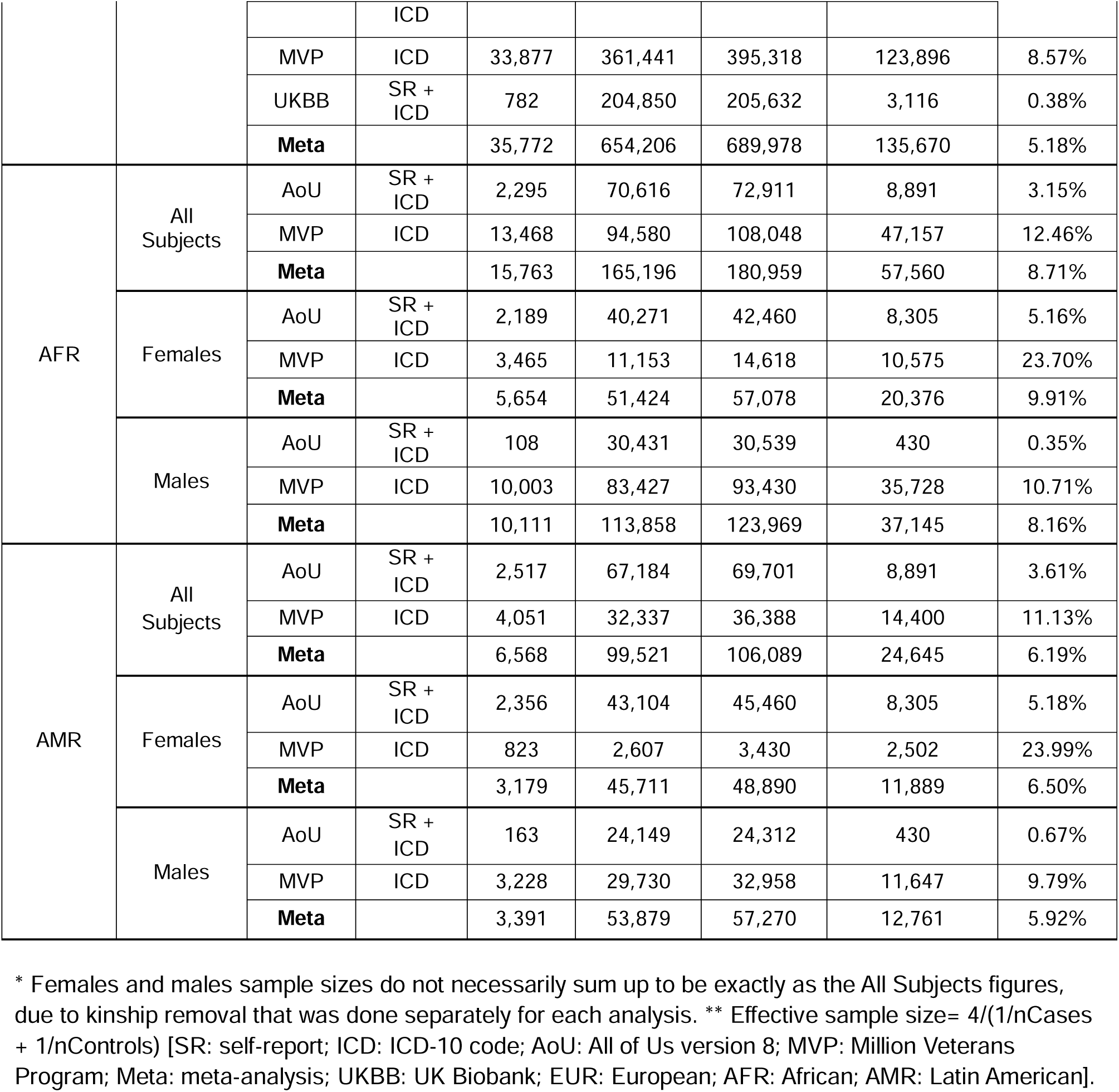
Demographics.

### Genome-Wide Association Studies (GWAS)

We conducted GWAS of fibromyalgia in three EUR cohorts – defined by stringent ICD-10 code, less-stringent ICD-10 code and/or self-report in AoU, by stringent ICD-10 code and less-stringent ICD-10 code in MVP and by stringent ICD-10 code and self-report in UKBB – and found one lead variant in each, all different. In an EUR meta-analysis (n_effective_=240,278), which included fibromyalgia summary statistics of the aforementioned AoU, MVP and UKBB analyses, plus summary statistics of stringent ICD-10 definition of fibromyalgia from Finngen, we discovered ten independent lead SNPs: *LOC105378797*\*rs1993709, *FBLN7*\*rs72831629, *TRAIP*\*rs71080556, *PRR16*\*rs56405820, *BAG6*\*rs2242656, *TSBP1-AS1*\*rs9279546, *PBX3*\*rs6478712, rs2587363, rs181388182 and rs11395028 (**Table 2**, **Figure 1a**). For AFR and AMR ancestries, GWAS analyses of fibromyalgia were possible in two cohorts – AoU and MVP. In AFR, there were no significant regions in AoU and one in MVP - *POLR1C*\*rs186798404 – which survived in the AFR meta-analysis (**Figure S1**). In AMR there were no significant effects in any of the analyses (**Figure S2**). A cross-ancestry meta-analysis, which included the three ancestral meta-analyses, revealed 12 significant lead variants: LOC105378797*rs10889947, *FBLN7*\*rs72831629, *CAMKV*\*rs2681780, *PRR16*\*rs190161089, *BAG6*\*rs2242656, *TSBP1-AS1*\*rs9279546, *UHRF1BP1*\*rs16894959, *POLR1C*\*rs186798404, rs12555516, rs10819064, *CNNM2*\*rs75970938 and rs9536401 (**Table 2**, **Figure 1b**). Regional Manhattan plots of all our lead SNPs in the various meta-analyses are presented in supplementary **Figures S9-S31**. We also conducted sex-stratified analyses, with no significant results for males in any of the individual and meta-analyzed AFR and AMR cohorts. In EUR, in UKBB we found one GWS locus in *SUCLG2-DT*\*rs10539712, which did not survive the meta-analysis. In females, in EUR we found one significant lead SNP in UKBB: *CRYBG3*\*rs188327717; one in AoU: rs1030125981; and none in MVP. In the EUR meta-analysis we found three lead SNPs: *IP6K1*\*rs763622663, *PRR16*\*rs62381083 and *PCLO*\*rs73167394. We did not find any female-specific effects in AFR and AMR. (**Figures S3-S8**).

**Figure 1:**
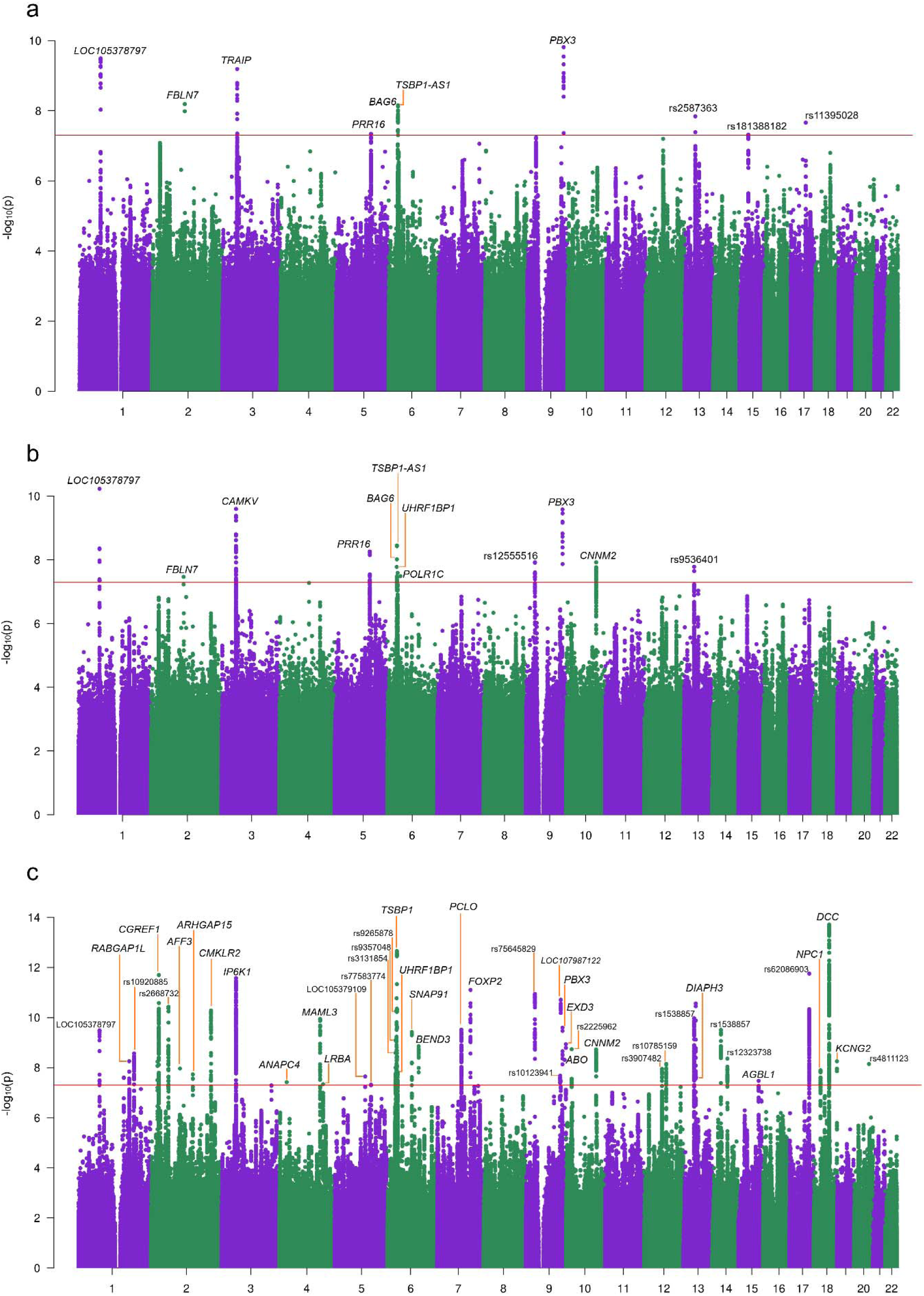
GWAS meta-analysis of fibromyalgia in **(a)** EUR population (n_total_=1,440,524, n_eff_=240,278), and **(b)** cross-ancestry (EUR-AFR-AMR; n_total_=1,727,572, n_eff_=323,773); **(c)** MTAG of fibromyalgia leveraging pain data (n_eff_= 240,278). Significant variants that are located within or near a gene are annotated according to the relevant gene; significant variants which are not located in the proximity of a gene are annotated with their rsID.

**Table 2:**
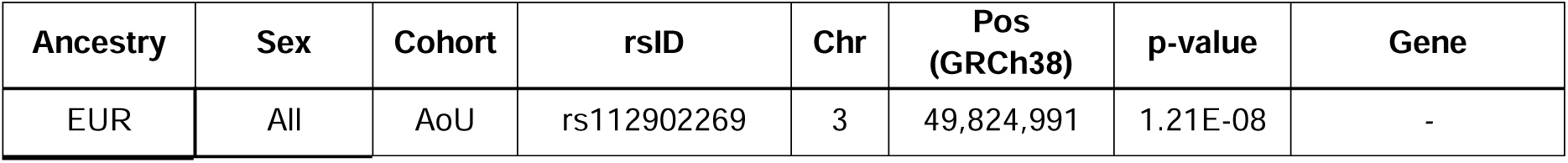
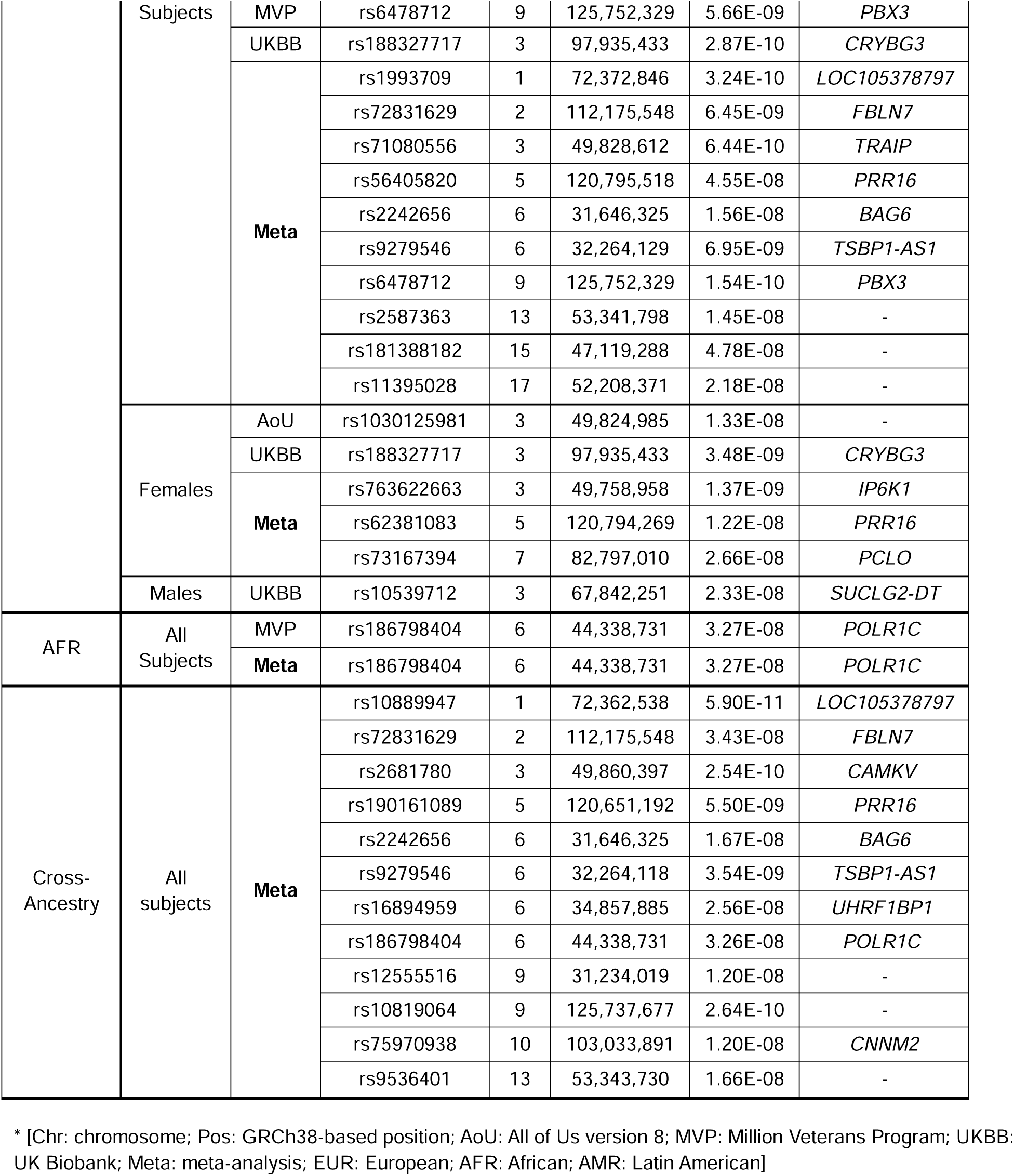
GWAS Lead SNPs.

### Gene-based and gene set analyses

Using MAGMA gene-based analysis we found 147 genes significantly associated with fibromyalgia in EUR. The strongest effects were for *PCLO*, *RNF123* and *DCC* (**Table S3**). In AFR, we found one gene associated with fibromyalgia: *TEX22* (**Table S4**). In AMR, we found no significant genes associated with fibromyalgia (**Table S5**). Cross-ancestry, we found 38 genes significantly associated with fibromyalgia (**Table S6**).

### Heritability Estimates and Genetic Correlations

We used LDSC to calculate the heritability estimates (h^2^) of the individual and meta-analyzed EUR cohorts, and found that the SNP heritability of fibromyalgia in the EUR meta-analysis is 6.9% (h^2^=0.069 ± 0.004) (**Table S7**). We also calculated inter-cohort (i.e., AoU, MVP, UKBB, Finngen) genetic correlations, and found significant values for all the pairs tested, with r_g_ values ranging between 0.72 (± 0.1) and 0.95 (± 0.1) (**Table S8**). We repeated these tests for the sex-stratified analyses (**Tables S7-8**). The genetic correlation between fibromyalgia in males vs females was r_g_=0.73 (± 0.064; p=3.7 × 10^−30^). We then calculated the genetic correlations between the EUR meta-analysis of fibromyalgia and 25 traits which are associated with cognitive function, physical activity, substance use, psychiatric disorders, autoimmune, cardiovascular and general health measures. All the r_g_ values were statistically significant. The strongest negative genetic correlation was with physical activity (r_g_ = −0.5 ± 0.027). The strongest positive genetic correlations were with chronic pain (r_g_ = 0.8 ± 0.023), PTSD (r_g_ = 0.72 ± 0.026) and depression (r_g_ = 0.69 ± 0.021), while the next three were of the PTSD subphenotypes hyperarousal (r_g_ = 0.68 ± 0.032), avoidance (r_g_ = 0.63 ± 0.034) and re-experiencing (r_g_ = 0.62 ± 0.032) (**Table S9**, **Figure 2**). For male-only and female-only analyses, the genetic correlations with all 25 traits were in the same direction as in the main analysis. All but four correlations in the male-only analysis were significant after Bonferroni correction (**Figure S32**, **Table S9**). We also repeated the genetic correlation calculation for all 25 traits using the fibromyalgia MTAG, with almost identical and all-significant results (**Figure S33**).

**Figure 2:**
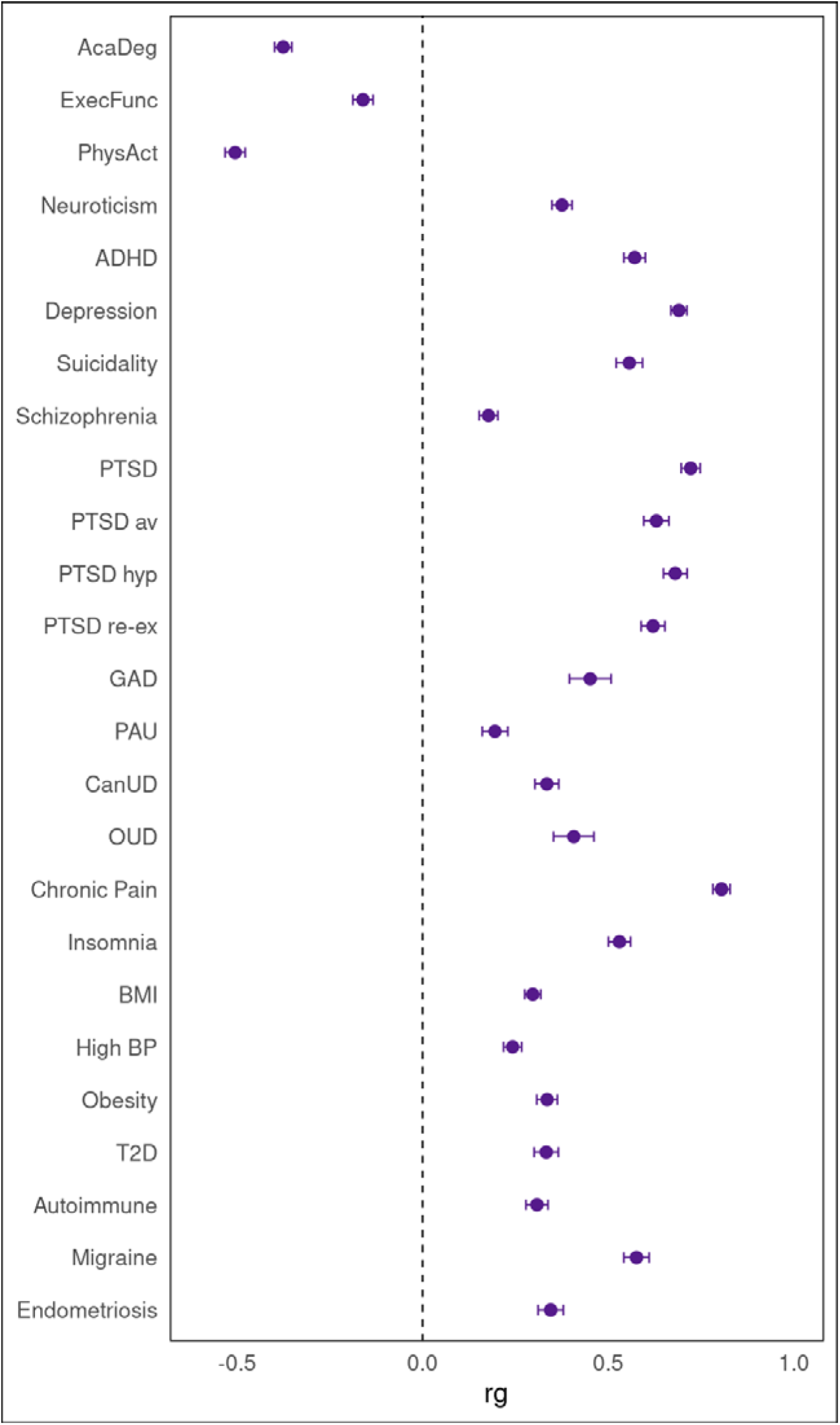
Genetic correlations between fibromyalgia and 25 in EUR, tested using LDSC. All the correlations are statistically significant [AcaDeg: academic degree; ExecFunc: executive functioning; PhysAct: physical activity; ADHD: attention-deficit/hyperactivity disorder; PTSD: post-traumatic stress disorder; av: avoidance; hyp: hyperarousal; re-ex: re-experiencing; GAD: generalized anxiety disorder; PAU: problematic alcohol use; CanUD: cannabis use disorder; OUD: opioid use disorder; BMI: body mass index; BP: blood pressure; T2D: type 2 diabetes].

### MTAG

MTAG was conducted using pain summary statistics [20] to increase the available information for the fibromyalgia GWAS results. The pain phenotype was a generalized trait that included diagnoses of limb, back, neck, head and abdominal pain The genetic correlation between fibromyalgia and pain was r_g_=0.72 (± 0.026; p=1.54 × 10^−169^). SNP-based heritability of the MTAG was 8% (h^2^=0.08 ± 0.003) (**Table S7**), and the genetic correlation between the fibromyalgia GWAS and MTAG (based on it but including also fibromyalgia variance retrieved from the pain GWAS) was r_g_ = 0.93 (± 0.007) (**Table S8**). The genetic correlations between the fibromyalgia MTAG and a variety of other traits were almost identical to the genetic correlations with the fibromyalgia GWAS (**Table S9, Figure S33**), indicating that the “enhanced” GWAS behaved similarly to the original GWAS in terms of its genetic relationships. The MTAG fibromyalgia GWAS resulted in 45 independent significant lead SNPs, an increase of 35 compared to the EUR fibromyalgia GWAS. Of the ten GWS loci in the fibromyalgia GWAS, seven were retained in MTAG filtering (that is, only seven of the ten original independent SNPs could be studied in the MTAG context; two were filtered out because they did not exist in the pain summary statistics, and one because it was multiallelic). The strongest effects in the MTAG were for rs62098042\**DCC* (p=1.95 × 10^−14^) and rs3129905\**TSBP1* (p=2.22 × 10^−13^) (**Figure 1c**, **Table S10**). We also conducted MTAG using both pain and LOO summary statistics of MDD together [47], excluding for the MDD data cohorts that overlap with those we used for the EUR fibromyalgia GWAS. The genetic correlation between fibromyalgia and the LOO-MDD data was r_g_=0.63 (± 0.025; p=1.05 × 10^−141^), and between pain and LOO-MDD was r_g_=0.56 (± 0.021; p=3.18 × 10^−150^). MTAG resulted in 94 independent significant lead SNPs, 34 of which were significant in the pain-leveraged MTAG (**Table S11**).

### MR

We used MRlap to assess the inferred causality between genetic liability to fibromyalgia and genetic liability to the traits included in the LDSC analyses. We used a p-value threshold of 1×10^−5^ to define genetic instruments. We found significant causal effects of fibromyalgia (as exposure) on 22 of the 25 traits we examined and a significant causal effect of 21 traits on fibromyalgia (as outcome). All the effects in this test were in the same direction as in the genetic correlations analysis. Problematic alcohol use (PAU) was the only trait which had no causal correlation with fibromyalgia in either direction (**Figure 3**, **Tables S12-13**) (an MR analysis with a p-value threshold of 1×10^−8^ was conducted as well; results are presented in **Table S14-15**).

**Figure 3:**
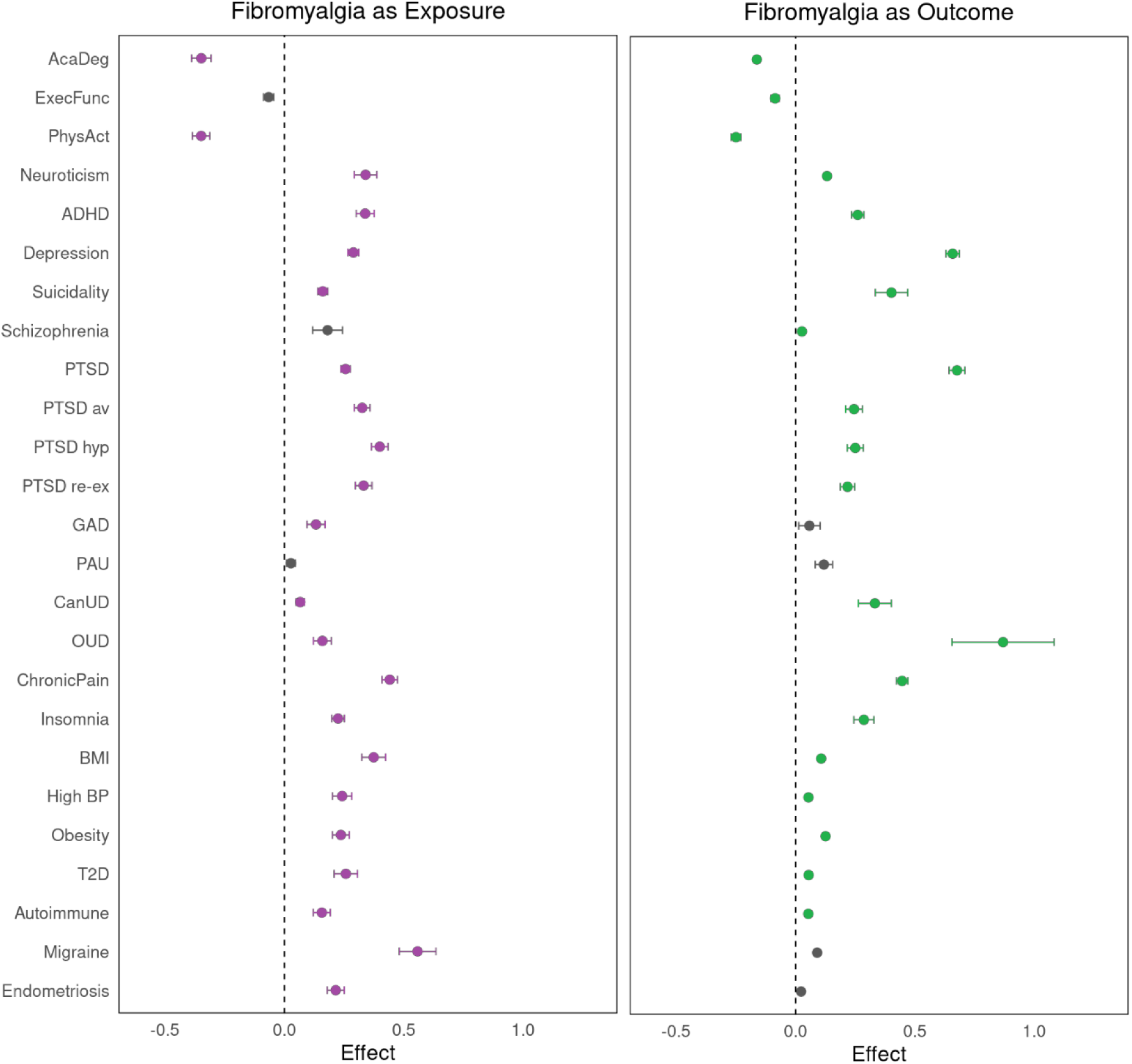
Causal genetic correlations between fibromyalgia and 25 traits in EUR, tested using MRlap. Non-significant values are plotted in gray [AcaDeg: academic degree; ExecFunc: executive functioning; PhysAct: physical activity; ADHD: attention-deficit/hyperactivity disorder; PTSD: post-traumatic stress disorder; av: avoidance; hyp: hyperarousal; re-ex: re-experiencing; GAD: generalized anxiety disorder; PAU: problematic alcohol use; CanUD: cannabis use disorder; OUD: opioid use disorder; BMI: body mass index; BP: blood pressure; T2D: type 2 diabetes].

### LAVA

We used LAVA to calculate the local genetic correlations between fibromyalgia and the 25 traits that were used to calculate LDSC in EUR (see above). Seventeen traits had at least one local genetic correlation with fibromyalgia – including all three PTSD subphenotypes – most prominently PTSD (seven loci) and chronic pain (5 loci). In total there were 45 local genetic correlations considering all pairs, involving 32 different genetic loci. The region between chr2: 59,024,862:60,547,931 correlated five different traits – chronic pain, depression PTSD and two of its subdimensions (hyperarousal, re-experiencing) – with fibromyalgia. The region between chr11:112,755,447:113,889,019 correlated four different traits – cannabis use disorder (CanUD), executive functioning, neuroticism and PTSD – with fibromyalgia (**Table S16**).

### TWAS

We used TWAS to evaluate predicted changes in differential gene expression in EUR. We identified four independent associated genes in four different tissues: *DPYSL5* with a positive enrichment in the cerebellum, and *DAG1*, *GPX1* and *PBX3* with negative enrichment in the tibial artery, skeletal muscles and pancreas, respectively (**Table S17**).

### gSEM

We performed gSEM to examine the overarching genetic relationships between fibromyalgia and 22 traits of interest that had demonstrated significant genetic correlations with fibromyalgia. EFA suggested a seven-factor model fit the data best, explaining 68.9% of cumulative variance. Factor 1 (SS loading: 3.09) explained 13.4% of the variance, factor 2 (SS loading: 3.03) explained 13.2%, factor 3 (SS loading: 2.75) explained 12%, factor 4 (SS loading: 2.32) explained 10%, factor 5 (SS loading: 1.81) explained 8%, factor 6 (SS loading: 1.77) explained 8%, and factor 7 (SS loading: 1.09) explained 5% of the overall variance. Traits with EFA loadings >0.25 were evaluated on the respective factors in CFA. CFA suggested that the seven-factor model fit the data well via traditional fit indices including a comparative fit index of 0.89 and a standardized root mean square residual (SRMR) of 0.054 (**Table S18**). Fibromyalgia co-loaded on factor 3 with insomnia, physical activity, and traits related to pain and autoimmune response. Depression and PTSD co-loaded on factor 1 with neuroticism and generalized anxiety disorder (GAD) and on factor 6 with ADHD and suicidality. Factor 6 had a strong genetic correlation (r_g_=0.75) with factor 3. Suicidality co-loaded on factor 2 with substance dependence traits (**Figure 4**).

**Figure 4:**
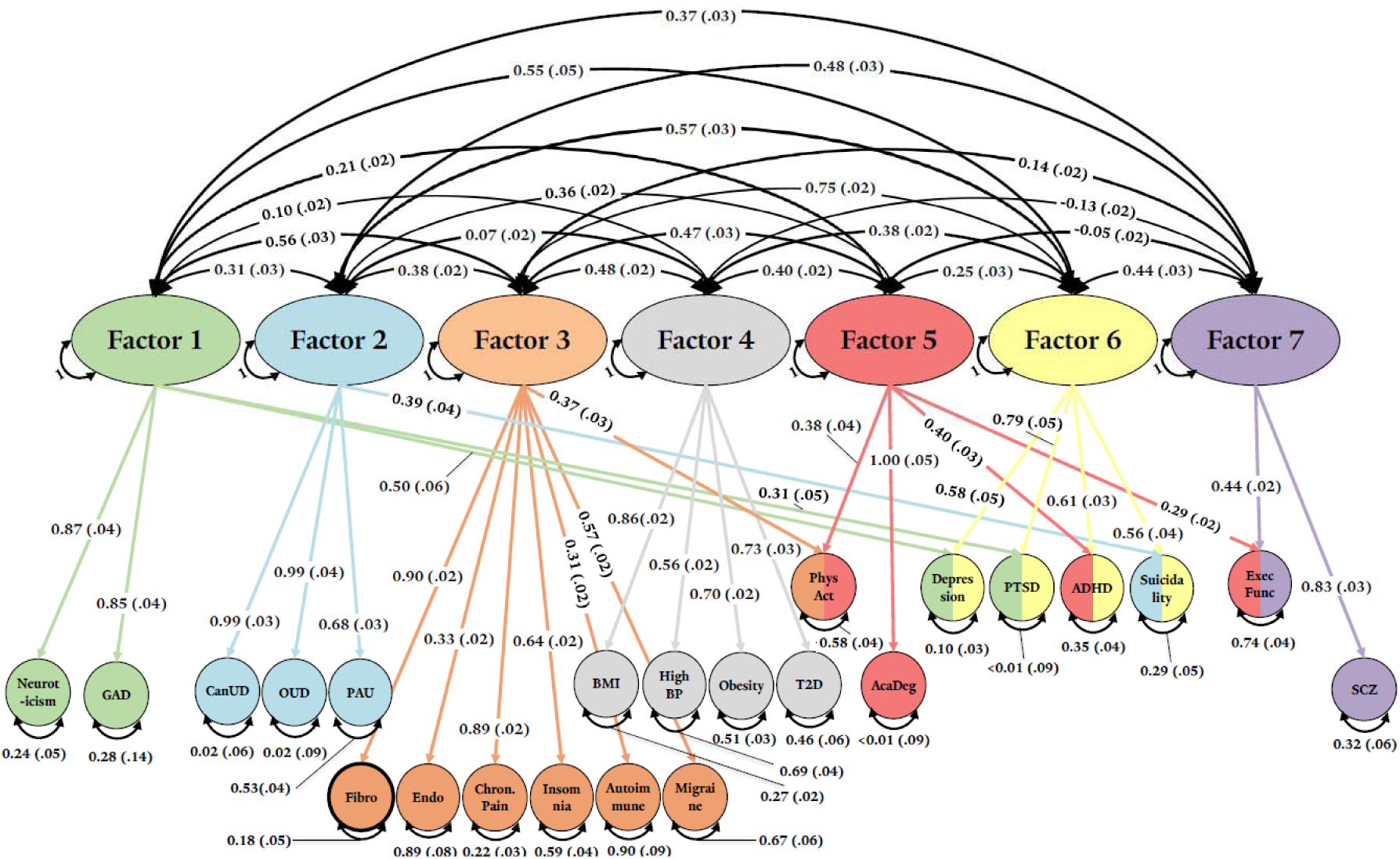
Genomic structural equation modeling (gSEM) assessing the overarching genetic relationship between fibromyalgia and 22 traits of interest. Fibromyalgia loaded on factor 3 [AcaDeg: academic degree; ExecFunc: executive functioning; PhysAct: physical activity; ADHD: attention-deficit/hyperactivity disorder; PTSD: post-traumatic stress disorder; av: avoidance; hyp: hyperarousal; re-ex: re-experiencing; GAD: generalized anxiety disorder; PAU: problematic alcohol use; CanUD: cannabis use disorder; OUD: opioid use disorder; BMI: body mass index; BP: blood pressure; T2D: type 2 diabetes; SCZ: schizophrenia].

### Drug Repurposing

After correction for FDR, we found no significantly enriched gene-drug pathways.

### Blood Type Phenotype

We find no differences between blood type distribution in fibromyalgia subjects compared to the general population (**Table S19**).

## Discussion

Fibromyalgia is a common syndrome that impairs the well-being of about 2.7% of the population (point prevalence). Existing knowledge regarding its etiology and potential treatments is sparse [2], especially considering its population-level impact. In this study, we identified ten independent GWS risk loci associated with fibromyalgia in EUR, one in AFR and twelve cross-ancestry. In an MTAG analysis that leveraged pain summary statistics, we found 45 independent loci associated with fibromyalgia. These findings, alongside genetic correlation, MR and gSEM analyses, provide a genotypic basis that supports the strong observed phenotypic associations between fibromyalgia and chronic pain, mental illnesses, autoimmune diseases, and unhealthy lifestyle as reflected by physical activity. Among the genetic variants that we identified, several map to genes that are associated with a variety of pathologies, protein expression and mental states, that can help explain the complex genetics behind the complex syndrome that is fibromyalgia:

*FBLN7* (fibulin 7; significant in the EUR and cross-ancestry meta-analyses) has a role in inflammation [56] and in anti-angiogenic activity [57]. It is also strongly associated with levels of MER proto-oncogene tyrosine kinase (MERTK) [58], a protein that is involved in retinal diseases [59] and in myelin phagocytosis in multiple-sclerosis (MS) patients [60]. MERTK is also involved in the pathology of systemic lupus erythematosus [61], an autoimmune disease that was previously genetically associated with fibromyalgia through a polygenic risk score (PRS)-based pheWAS [8], in line with a moderate genetic correlation (r_g_=0.31) we found between fibromyalgia and autoimmune diseases.

*PBX3*, significant in the EUR meta-analysis, is involved in the etiology of several types of cancer [62] and influences systolic blood pressure (BP) [63, 64]. It affects the structure of the brain basal ganglia [65], which play important roles in executive functioning and emotional regulation [66]. It is also associated with gastroesophageal reflux disease (GERD) [42], which was genetically associated with fibromyalgia through PRS [8]. In the TWAS, *PBX3* was found to be significantly enriched in the pancreas, which could indicate a potential role in the high rates of gastrointestinal disease and pancreatitis among fibromyalgia patients (4-10 times compared to the general population) [67].

In three of the GWAS we conducted - the EUR, the female-specific EUR and the cross-ancestry meta-analyses, there were three different lead variants on chromosome 3, which map to three different genes but may represent the same GWS signal. This assumption is based on FUMA outputs, which pointed to a single genomic risk locus at this region in each of the analyses (despite that all three are within a ∼100 kb region). Regional Manhattan plots of this locus visually demonstrate the proximity of these genes to each other and the distribution of significant variants across all three of them (**Figures S11, S22**). These genes are *TRAIP*, *IP6K1* and *CAMKV*, and they all have significant associations in the EUR and cross-ancestry gene-based analyses (MAGMA) too. *TRAIP* encodes the TRAF interacting protein and is important for DNA repair [68]. It is a strong determinant of the expression of TXNDC12, a protein involved in ferroptosis [69, 70], a cell death process triggered by lipid peroxidation, which may cause neuropathic pain [71] and increased fibromyalgia symptoms [72]. *TRAIP* is also genetically associated with ADHD [73], cognitive functioning [74], intelligence [75], insomnia [41] and pain intensity [76] – all are traits that we found to be genetically correlated with fibromyalgia, directly (ADHD, insomnia) or indirectly (e.g. academic degree is a proxy to intelligence, chronic pain and migraine are associated with pain intensity). *IP6K1* is associated with insomnia [41], cognitive ability [77], and with levels of MST1 [58], a protein that affects inflammation and autoimmune response [78]. *MST1*, as well as *MST1R* (which encodes the MST1 receptor), are closely located to *TRAIP* and *IP6K1* on chromosome 3 and both had a significant effect in the gene-based analysis. *RNF123*, which resides only 83 bp from *MST1*, adds another strong association with fibromyalgia in the EUR and cross-ancestral gene-based analyses; this gene was previously associated with chronic widespread musculoskeletal pain, a common symptom of fibromyalgia [10]. *CAMKV*, significant in our cross-ancestry meta-analysis, was previously shown to affect intelligence [79], educational attainment [80, 81] and the pleiotropy between BMI and osteoarthritis [82].

*TSBP1-AS1* – significant in the EUR and cross-ancestry analyses – has a strong effect on hypothyroidism and rheumatoid arthritis [42, 83], autoimmune disorders that share symptoms with fibromyalgia, like pain and muscle tenderness, in line with the genetic correlation between fibromyalgia and autoimmune diseases. *TSBP1-AS1* has also a robust effect on the levels of the immunomodulatory C2, C4, and LILRB4 [58, 84, 85], the deficiency of which is associated with obesity and type 2 diabetes (T2D) [86]. Both of these traits were moderately genetically correlated with fibromyalgia and may lead to lupus-like effects [87], further establishing the role of autoimmune response in fibromyalgia. In addition, obesity, BMI and autoimmune response are locally genetically correlated with fibromyalgia in loci that do not overlap but are adjacent (<1.5MB) to *TSBP1-AS1*. Also within this region, we found body mass index (BMI) to be locally genetically correlated with fibromyalgia in a locus that includes *SNRPC* - significant in the EUR gene-based analysis, and previously associated with lupus erythematosus [88] and with smoking initiation [89]. *BAG6*, closely located to *TSBP1-AS1* on chromosome 6 and significant in our GWAS, is associated with the autoimmune condition psoriasis [90], which may cause joint pain and arthritis.

We conducted an MTAG analysis, leveraging pain summary statistics to create a better-powered genome-wide analysis of fibromyalgia. High r_g_ between the GWAS and MTAG, and almost identical genetic correlations with other traits, suggest that the MTAG results indeed represent the genetics of fibromyalgia. Out of 28 MTAG GWS loci in gene-coding regions, 15 had a significant effect in at least one of the EUR or cross-ancestry GWAS or MAGMA analyses, further confirming the MTAG validity. *PCLO*, significant in the MTAG and in the female EUR GWAS, had the strongest effect in the EUR gene-based analysis. It affects bone mineral density (BMD) [91], which is associated with fibromyalgia symptoms (i.e., BMD is lower in fibromyalgia subjects) [92], as is *DCC [91]*, which had the strongest effect in the MTAG, and is also associated with smoking initiation [89], educational attainment [93], neuroticism [94], and various types of pain [20]. *MAML3* and *FOXP2* were also previously associated with pain intensity [76]. Another MTAG-GWS gene of interest is *ABO*. A blood type (ABO blood group)-determining protein, *ABO* has a major impact on a variety of blood protein expression phenotypes [84, 95]. Several studies point to an interaction between blood type and pain perception [54, 55]. However, we did not find any phenotypic differences in blood type distribution among fibromyalgia patients compared to the general population.

Several other genes that were GWS in our GWAS and/or MTAG will be discussed briefly: *UHRF1BP* and *ARHGAP15* are associated with BMD [91]. *ARHGAP15* was also found to be the strongest genetic factor contributing to diverticular disease [96] and was associated with educational attainment, as were *KCNG2*, *CNNM2*, *EXD3* and *AFF3* [93]. *AFF3* also affects rheumatoid arthritis [97], and is part of a list of genes that affect substance use traits such as smoking initiation and alcohol consumption, along with *SNAP91*, *CNNM2*, *NPC1* and *RABGAP1L* [89]. Though not mapping to a coding gene, rs1993709 was associated with BMI, obesity and weight [98, 99] and also with neuroticism [100]. Data obtained from GTEx portal (gtexportal.org) [101] indicated that rs1993709 is a very strong negative eQTL of the pseudogene *RPL31P12* transcription in the cerebellum (normalized effect size = −0.87; p=2.7×10^−16^). Though its function is not clear, *RPL31P12* had a significant effect in studies of BMI, weight and obesity [20]. The non-coding rs2587363 was previously associated with pain [9].

Genetic correlation analysis revealed moderate-to-strong positive associations between fibromyalgia and several psychiatric traits, namely depression, suicidality, ADHD and PTSD, including the three DSMIV PTSD subphenotypes of reexperiencing, avoidance, and hyperarousal. These correlations are in line with high comorbidity between fibromyalgia and these traits [11–13]. In people with both PTSD and fibromyalgia, the onset of fibromyalgia symptoms is usually later than the occurrence of the traumatic event, [102] a time course consistent with possible causality. While MR revealed bidirectional causality between these traits, the causal effect of PTSD (as exposure) on fibromyalgia (as outcome) was much stronger (although not for the subphenotypes of PTSD), suggesting that the genetic liability to PTSD may be more likely to cause fibromyalgia than vice versa. The same is true regarding depression and suicidality, but not ADHD: MR results show that genetic liability to fibromyalgia is more likely to cause ADHD; it is possible that what is reflected here are concentration and attention problems (including what is sometimes referred to as “brain fog”) --- known to be common in fibromyalgia -- that might be coded as ADHD. The moderate r_g_ between fibromyalgia and neuroticism aligns with a known genetic correlation between neuroticism and various pain-related traits [40, 103, 104], and with dominant neuroticism in fibromyalgia patients compared to other personality traits of the Big-Five model, as was found in a personality-trait questionnaire conducted among fibromyalgia patients [105].

The psychiatric traits described above are strongly associated with chronic pain [40, 104], one of the most dominant symptoms of fibromyalgia. Chronic pain was the trait most strongly genetically correlated with fibromyalgia in our study (r_g_ = 0.80 ± 0.02), alongside a relatively high correlation with migraine - a more specific pain-related trait. In a gSEM analysis, fibromyalgia co-loaded with chronic pain and migraine, as well as with endometriosis, autoimmune response, insomnia, and physical activity, but not with psychiatric traits. This suggests that fibromyalgia may be contextualized more as a pain and autoimmune trait than a psychiatric trait, even though the genetic correlations with psychiatric traits such as PTSD and depression are quite high. Genetic correlations between fibromyalgia and opioid use disorder (OUD; r_g_ = 0.41 ± 0.05) and CanUD (r_g_ = 0.33 ± 0.03) are of importance here, due to the use of opioids to alleviate pain, and the growing use of cannabis in recent years to treat symptoms of fibromyalgia [2, 14, 106, 107]; the moderate genetic risk of developing dependence of these substances among fibromyalgia patients should be taken into account when prescribing them as treatment. MR analysis revealed that fibromyalgia has a weak causal (yet statistically significant) effect on CanUD and OUD risk.

A moderate negative genetic correlation between fibromyalgia and physical activity (r_g_ = −0.5 ± 0.03) reinforces the importance of physical activity as a possible treatment for fibromyalgia symptoms [2] (alternatively, that fibromyalgia itself causes reduction in physical activity, as seen in the bidirectional results in the MR analysis). Along with a positive genetic correlation we found between fibromyalgia and BMI, obesity, high BP and T2D, our observations provide additional support for a biological connection between a healthy lifestyle and fibromyalgia. The moderate r_g_ fibromyalgia had with endometriosis is also backed by the strong phenotypic association between these traits [108]. Negative genetic correlations between fibromyalgia and executive functioning and with academic degree indicate a genetic relationship underlying the phenotypic association between fibromyalgia and impaired cognitive performance [109]. The moderate genetic correlation we identified between fibromyalgia and autoimmune diseases (r_g_ = 0.31 ± 0.03), along with the autoimmune roles of several GWS genes in our study, point to the possibility that fibromyalgia has autoimmune components. This notion was suggested previously [110], with studies linking fibromyalgia and autoimmune diseases through genetic and inflammatory pathways [8, 111].

LAVA analysis revealed local genetic correlations between fibromyalgia and several traits in regions that map to genes of interest. Four traits – executive functioning (negative r_g_ value), CanUD, neuroticism and PTSD (positive r_g_) – were correlated with fibromyalgia in a region on chromosome 11 that maps to *TTC12*, which had a significant effect in the EUR gene-based analysis. This gene affects neuroticism [112], alcohol dependence [113], and nicotine dependence traits [114, 115], and is an important factor in the expression of NCAM1, a cell adhesion molecule that is genetically associated with a variety of substance use traits [89, 114–116]. It also is physically adjacent to *DRD2*, which encodes the dopamine D2 receptor and is associated with psychiatric and behavioral traits such as MDD [35], anxiety [117] neuroticism [94], and a variety of substance use traits [39, 89, 118]*. NRXN1*, which maps to a region that genetically correlates fibromyalgia with academic degree, was previously associated with educational attainment [93]. *GRIA1*, which encodes the GluA1 subunit of the AMPA receptor, is associated with insomnia [41] and personality disorders [20], and it also maps to a region that genetically correlates fibromyalgia with PTSD. Administration of the cannabis-derivatives Δ9-tetrahydrocannabinol (THC) and cannabidiol (CBD) induced alterations in *Gria1* expression in rodents [119–121], consistent with growing evidence of cannabinoid-glutamatergic associations [122], suggesting a potential pathway through which cannabis may ameliorate fibromyalgia symptoms [106, 107].

Nine genes identified in the gene-based analysis map to a region on chromosome 2 that is locally positively genetically correlated with fibromyalgia and depression (there is a possible signal at the same region, which can be seen in **Figure 1a**, though there are no GWS variants). Four of these genes (*DPYSL5*, *MAPRE3*, *AGBL5*, *CGREF1*) are associated with blood triglyceride levels [58, 123, 124], two (*CENPA*, *SLC35F6*) with BMD [91], and two (*MAPRE3*, *DPYSL5*) with brain morphology [125, 126]. Another gene, *KHK*, encodes an enzyme with a major role in fructose metabolism.

This study included fibromyalgia data from four large biobanks. In AoU, UKBB and Finngen, heritability estimates were between 11-19%, and females were 82-92% of the total number of cases. In MVP, however, heritability was considerably lower (3%), and females accounted for only 19% of the fibromyalgia cases (compared to 7% of the sample). Nevertheless, the incidence of fibromyalgia in MVP was much higher, compared to the other cohorts we investigated (**Table 1**). While the different case distribution among sexes in MVP is attributable to the sex imbalance in the cohort, the low heritability is puzzling and may relate to the military nature of MVP. Indeed, soldiers tend to be more physically active in their daily routine compared to civilian population [127]. As MVP subjects are veterans, there may be higher likelihood that they were exposed to environmental factors that contributed to fibromyalgia symptoms during their service (e.g., injuries, traumatizing events). These two factors may explain the high prevalence but low genetic heritability of fibromyalgia among MVP participants. Nevertheless, genetic correlations between the EUR fibromyalgia cohorts were high (r_g_ values ranged between 0.72-0.95; **Table S8**), indicating that the fibromyalgia phenotype in all the cohorts we analyzed likely represents the same trait. A strong genetic correlation (r_g_=0.73 ± 0.06), between male and female EUR participants, as well as great similarities between the genetic correlations of males’ and females’ fibromyalgia datasets with other traits (**Table S9**; **Figure S32**), indicates that fibromyalgia likely represents the same trait in both sexes.

Our GWAS analyses yielded a total of 15 genomic loci associated with fibromyalgia across different ancestries, ten of which map to a coding gene – and the MTAG resulted in 36 additional GWS loci, 22 of which map to a coding gene. Several of these genes are associated with traits that share with fibromyalgia symptomatic characteristics like pain, muscle tenderness and insomnia, while others are associated with physical measures of general health like BP and BMI. Some of the genes we identified in this study are associated with structural changes in brain regions that mediate emotional regulation and executive functioning, with mental and cognitive phenotypes such as ADHD, educational attainment and cognitive functioning, and with substance use traits. In addition, at least three of the genes we found are associated with autoimmune disorders. These findings are all in line with moderate genetic correlations observed between fibromyalgia and phenotypes of general health (physical activity, T2D, high BP, BMI, obesity) and of substance use (CanUD, PAU, OUD), and moderate-to-strong genetic correlations of fibromyalgia with chronic pain, with psychiatric traits (ADHD, depression, suicidality, PTSD), and with autoimmune traits. Our findings may reflect that fibromyalgia is a combination of several factors that create a complex trait, which can be categorized under several different factors that contribute to its phenotypic expression, with gSEM suggesting that it may be considered mainly as pain- and autoimmune-associated.

This study has limitations. First, in two of the three cohorts in which we conducted a sex-stratified GWAS, the number of male participants with fibromyalgia (i.e., cases) was low and we did not detect GWS loci. Nevertheless, as discussed earlier, the genetic correlation between male and female fibromyalgia datasets was high, suggesting that the trait is similar or identical between the sexes. Second, only one GWS locus was detected in the AFR meta-analysis, and none in AMR, probably due to low sample sizes. Third, fibromyalgia prevalence differs between geographic regions and populations (for example: 9.3% in Tunisia, 8.8% in Turkey and 0.4% in Greece, compared to the world prevalence of 2.7% [128]) and also between different studies of the same region, suggesting that the true prevalence of this disorder in the population can be biased due to practices of diagnosis and environmental-cultural differences (in diet and exercise, as well as in symptom expression). Nevertheless, it may also represent actual genetic differences between populations that are underrepresented in large genetic biobanks, an issue that needs to be addressed in future studies, when more data regarding these populations are available.

In total, our results provide novel insights into the genetic architecture of fibromyalgia, providing novel information about its genetic association with pain, autoimmune response, psychiatric traits, and lifestyle. These results allow for a deeper understanding of fibromyalgia and may provide tools for future identification of genetic risk factors that affect this complex disorder, which in turn may help with diagnosis, treatment and even prevention of symptoms, with adoption of a healthier lifestyle, and other treatments that may arise in the future. We studied multiple populations. While we identified the greatest number of findings in EUR, there was also independent GWS in AFR. All populations contributed to increasing findings in our trans-population meta-analysis. There is future potential for medication repurposing and personalization of treatments, but considering the lack for results for the former in the present analyses, this will require future studies with more subjects and better power.

## Supporting information

Supplementary Figures and Acknowledgements

## Data Availability

All data will be made available upon publication

## Acknowledgements

This research is based in part on data from the Million Veteran Program, Office of Research and Development, Veterans Health Administration, and was supported by MVP000 as well as 5IO1CX001849. More details regarding this consortium are provided in the Supplementary Information. This publication does not represent the views of the Department of Veterans Affairs or the United States Government. This research has also been conducted using the UK Biobank Resource under Application Number 41910. We acknowledge All of Us participants for their contributions, without whom this research would not have been possible. We also thank the National Institutes of Health’s All of Us Research Program for making available the participant data examined in this study (researchallofus.org). Genotype-tissue expression data used for the analyses described in this manuscript were obtained from the GTEx Portal on 04/25/2025.

## Data Sharing and Availability

MVP summary data will be made available through dbGAP accession number phs001672.

