## Supplementary Figures and Acknowledgements for "The Genetics of Fibromyalgia and its Relationships to Psychiatric and Medical Traits"

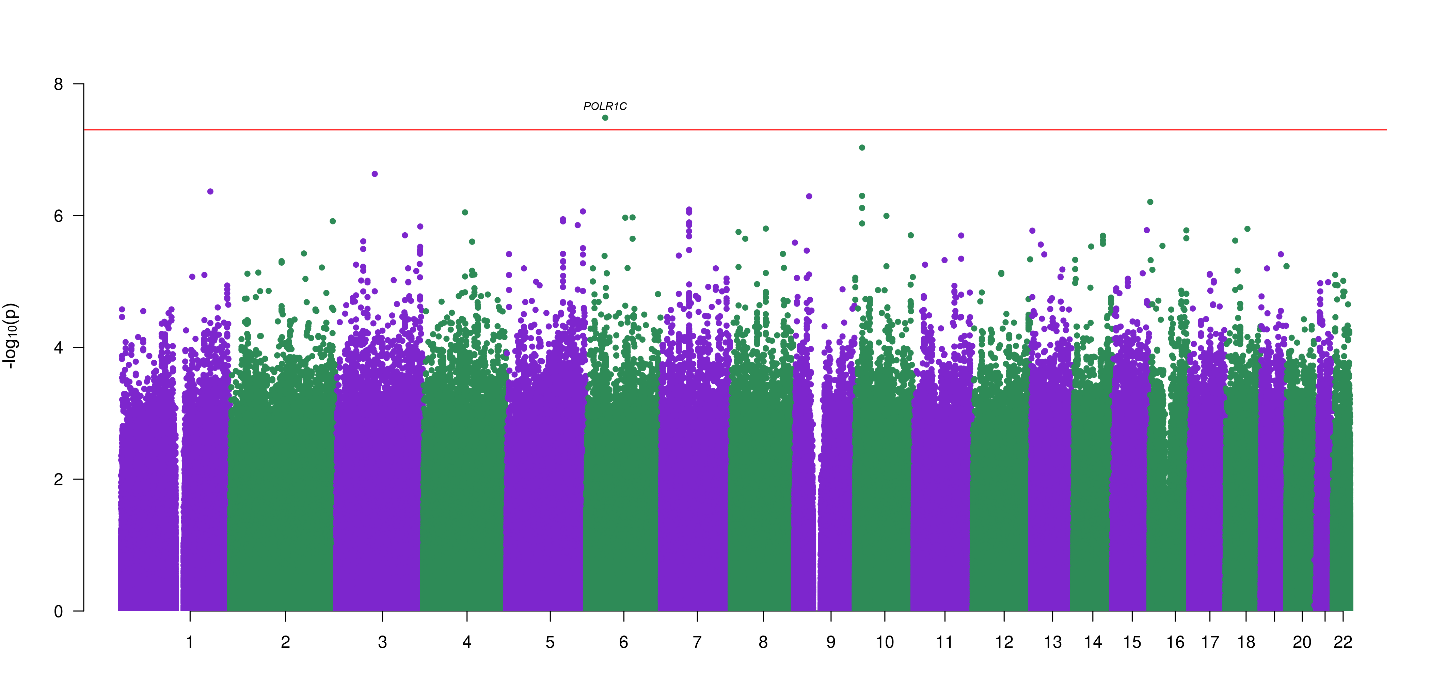

**Figure S1**: GWAS meta-analysis of fibromyalgia in AFR population (n_total_=180,959, n_eff_=57,560).

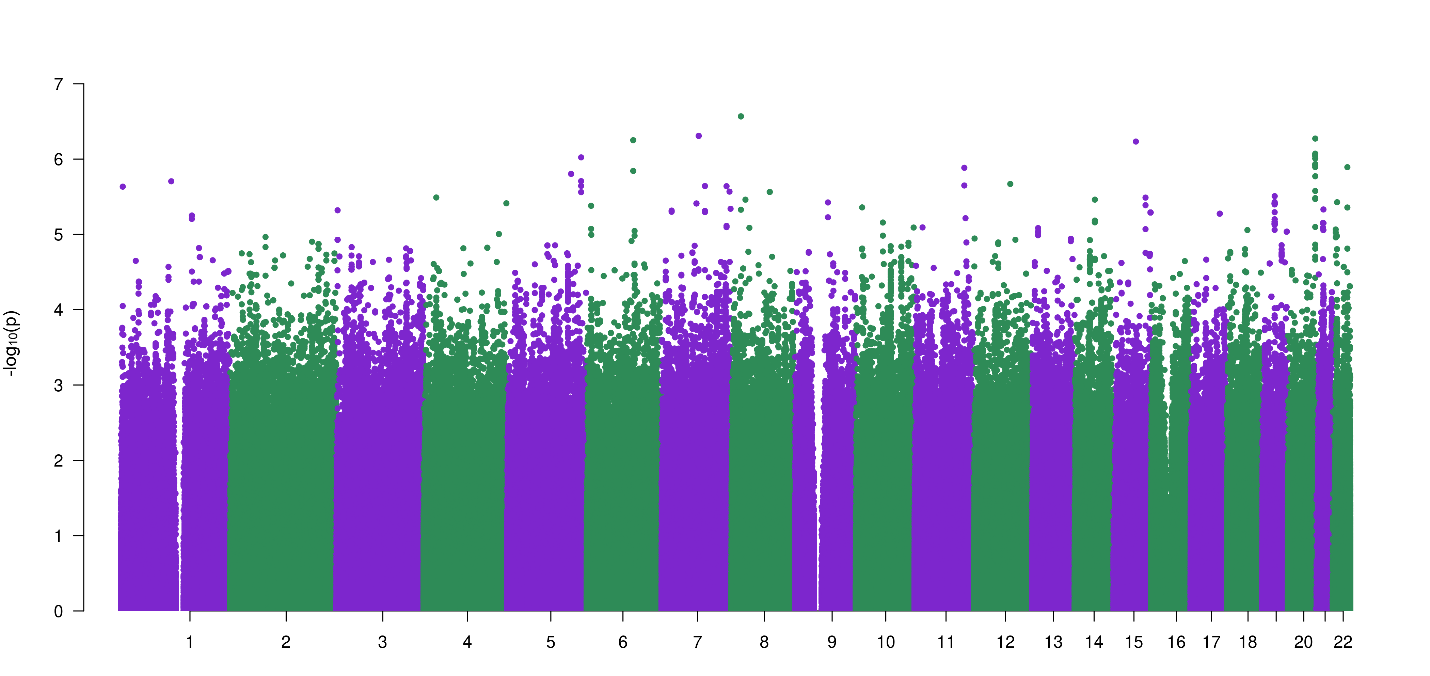

**Figure S2**: GWAS meta-analysis of fibromyalgia in AMR population (n_total_=106,089, n_eff_=24,645).

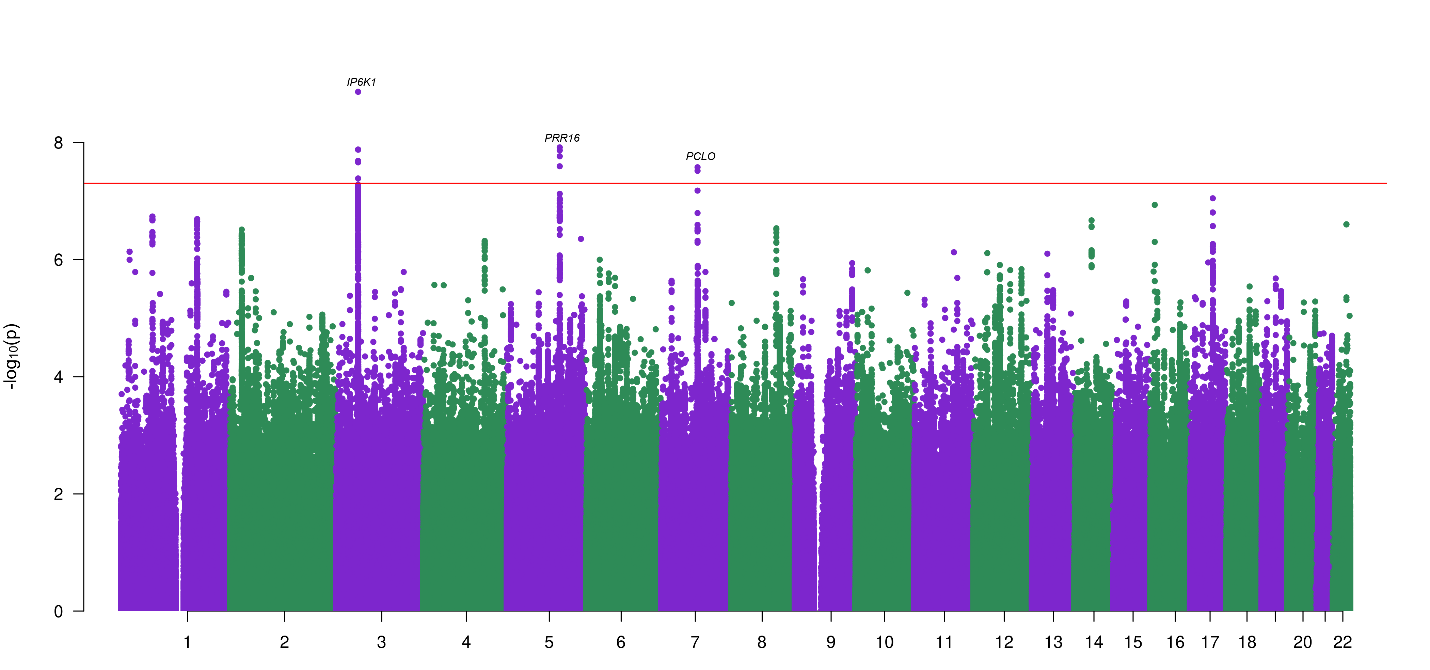

**Figure S3**: GWAS meta-analysis of fibromyalgia in EUR population - females (n_total_=401,602, n_eff_=87,471).

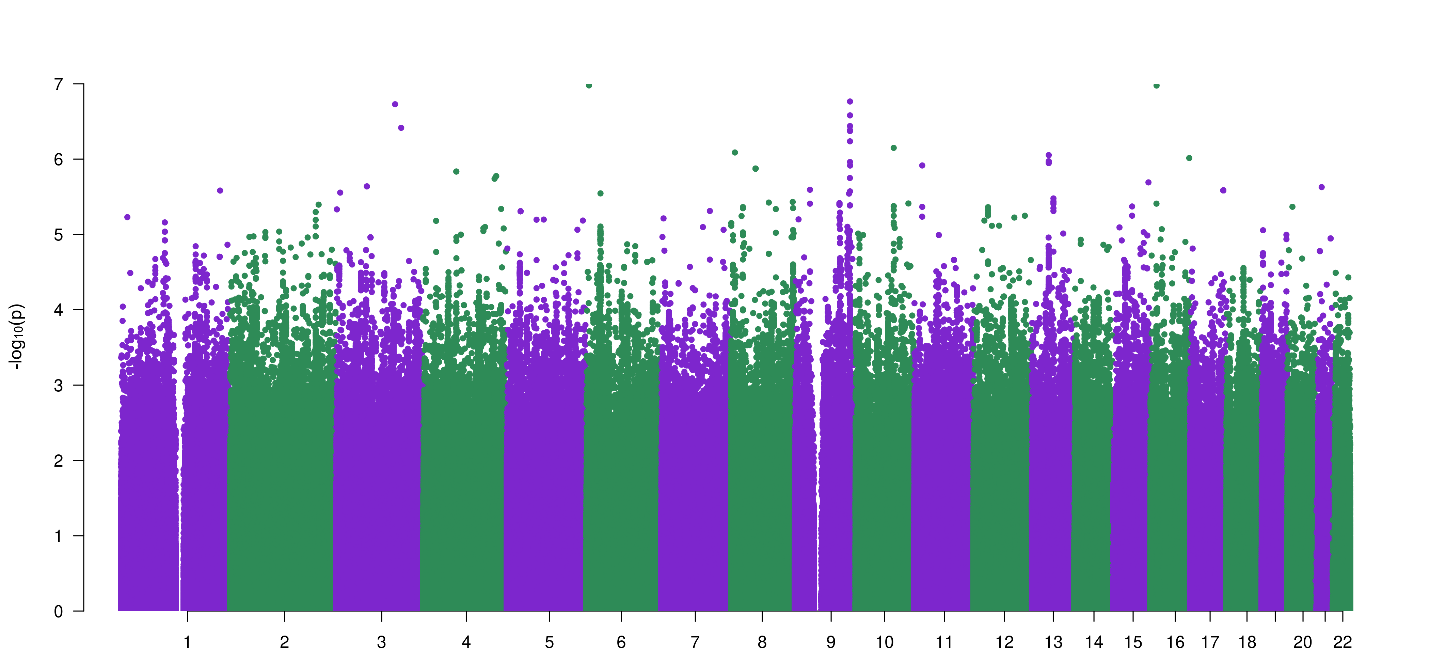

**Figure S4**: GWAS meta-analysis of fibromyalgia in EUR population - males (n_total_=689,978, n_eff_=135,670).

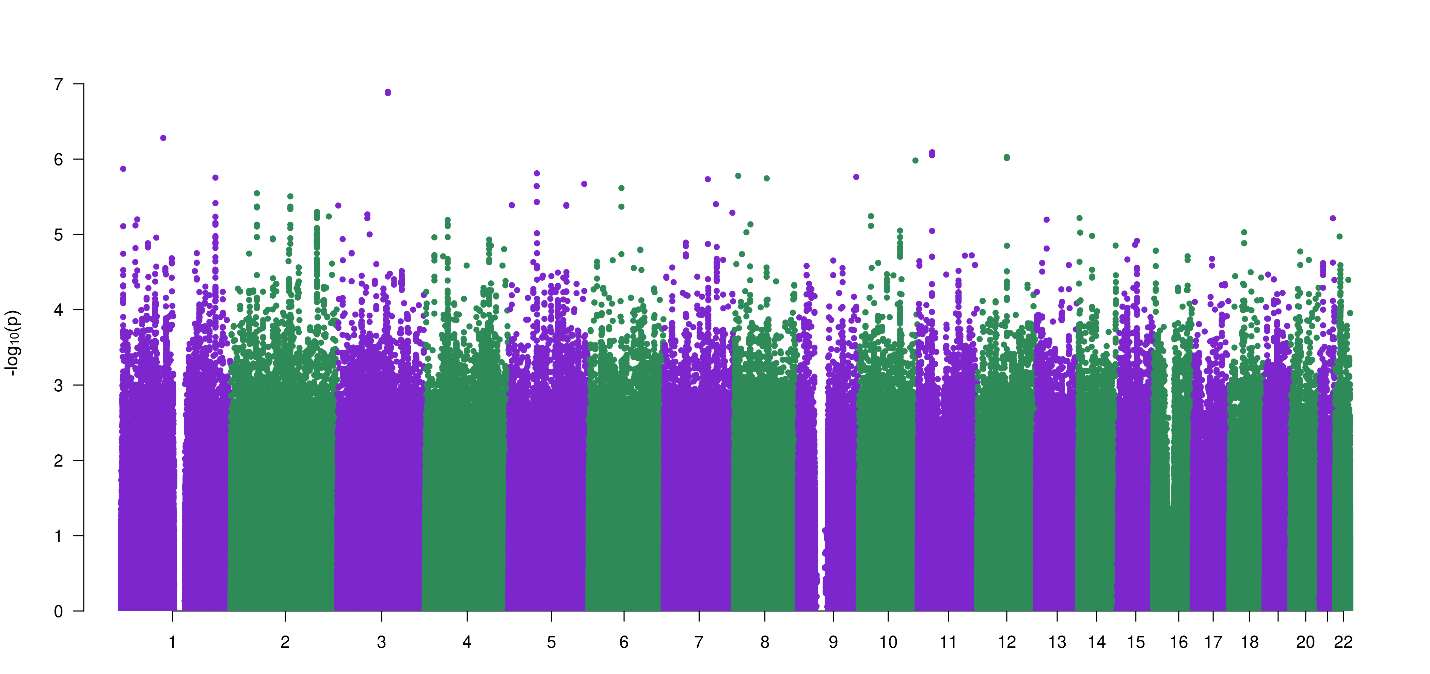

**Figure S5**: GWAS meta-analysis of fibromyalgia in AFR population - females (n_total_=57,078, n_eff_=20,376).

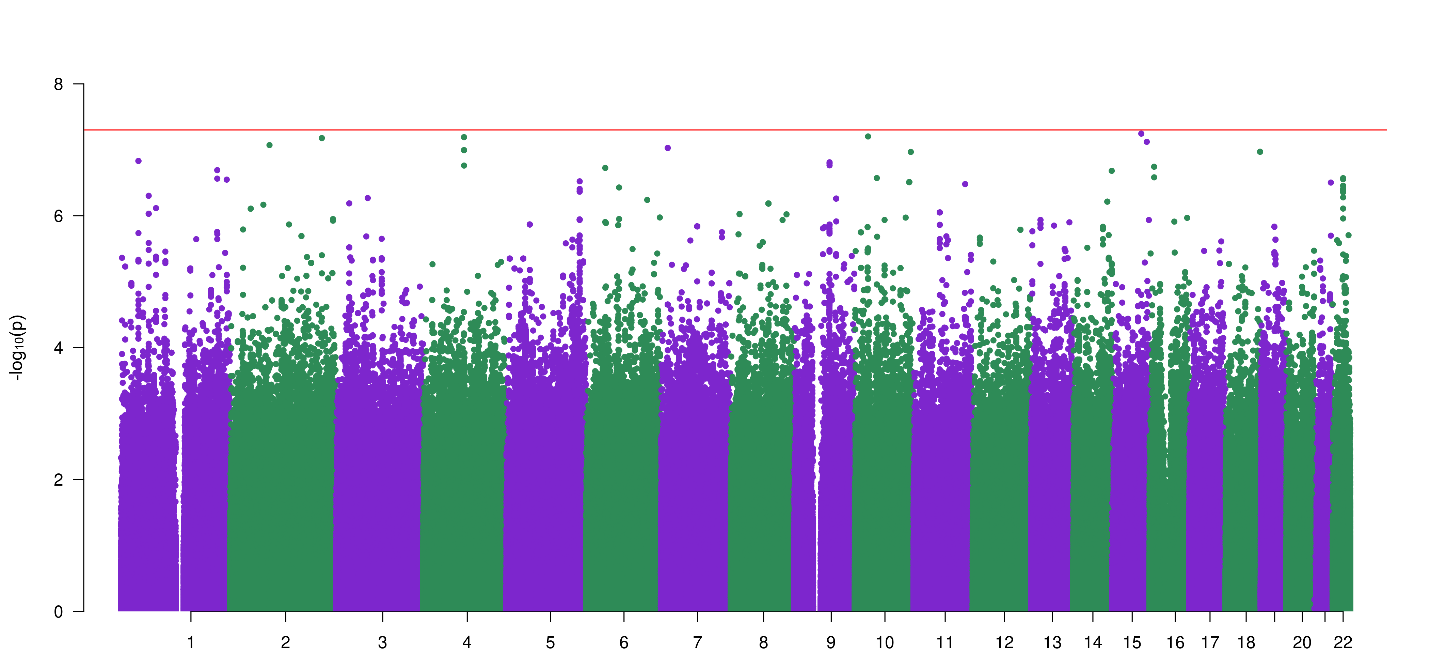

**Figure S6**: GWAS meta-analysis of fibromyalgia in AFR population - males (n_total_=123,969, n_eff_=37,145).

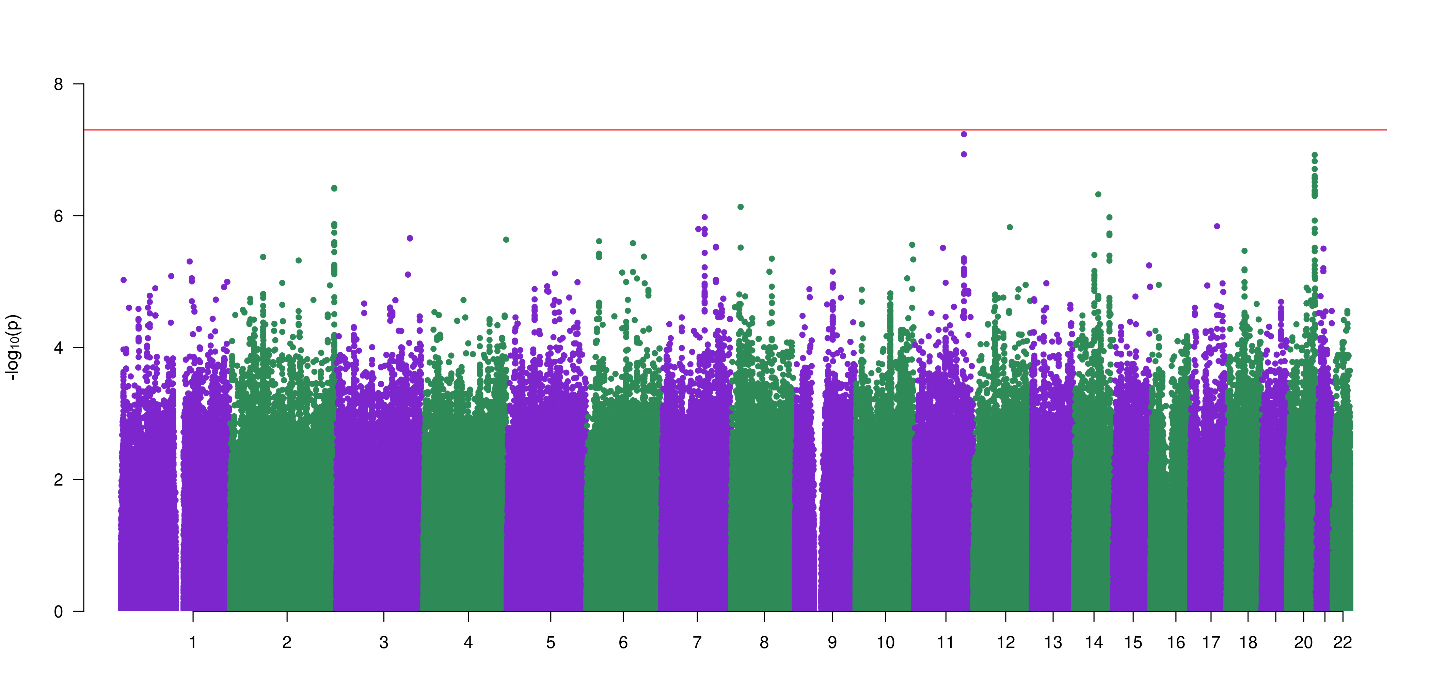

**Figure S7**: GWAS meta-analysis of fibromyalgia in AMR population - females (n_total_=48,890, n_eff_=11,889).

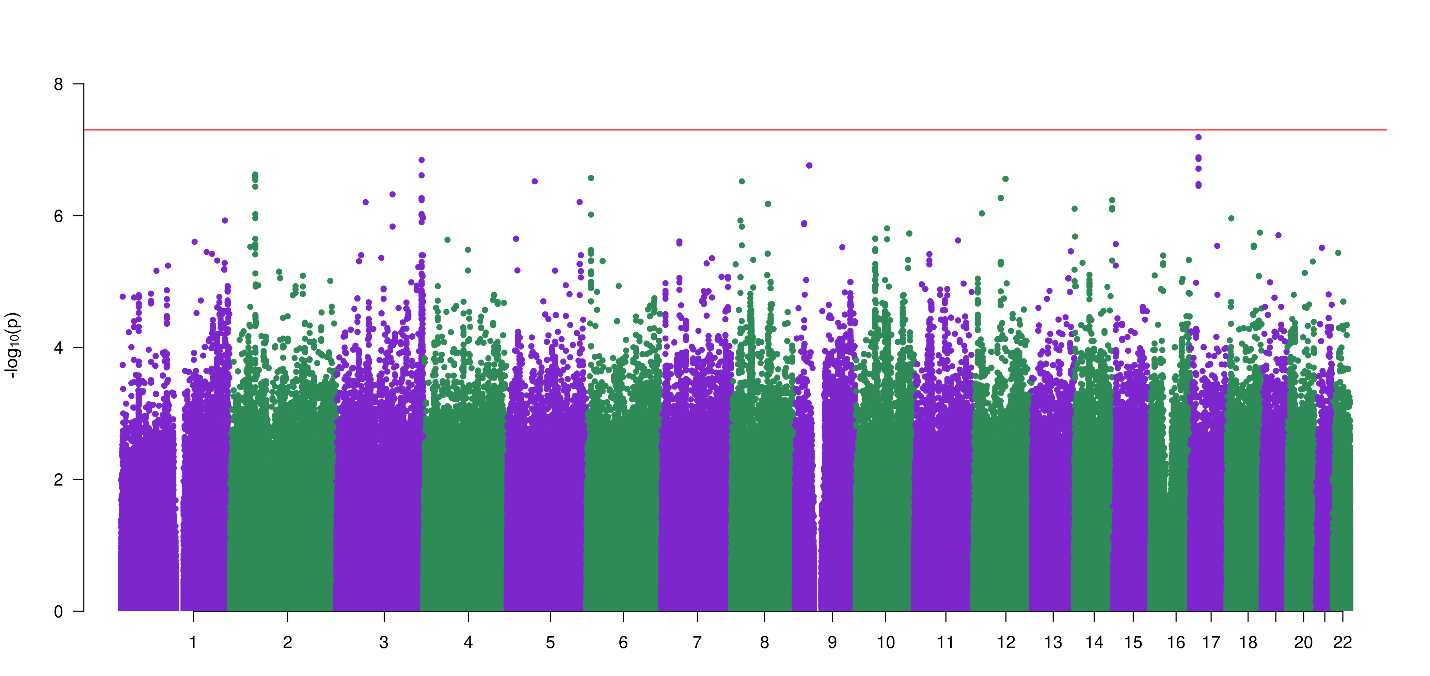

**Figure S8:** GWAS meta-analysis of fibromyalgia in AMR population - males (n_total_=57,270, n_eff_=12,761).

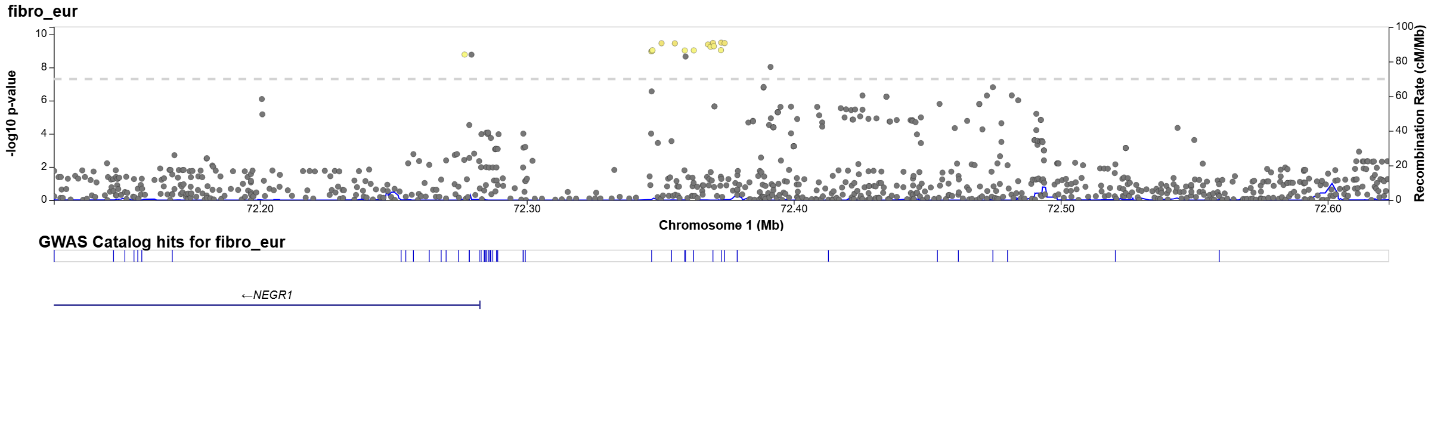
**Figure S9**: regional Manhattan plot: rs1993709 (EUR meta-analysis).

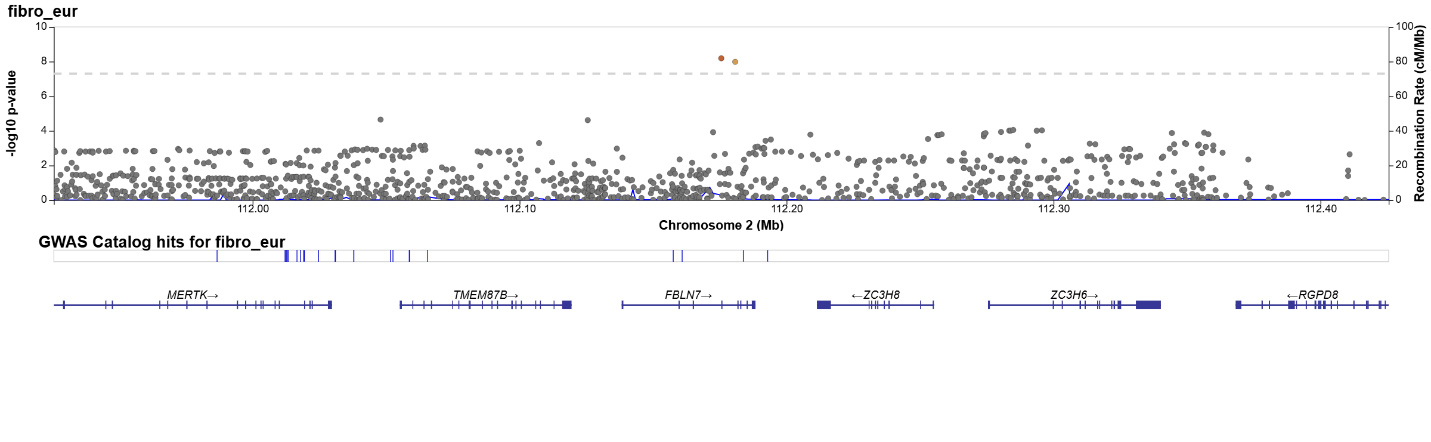
**Figure S10**: regional Manhattan plot: rs72831629 (EUR meta-analysis).

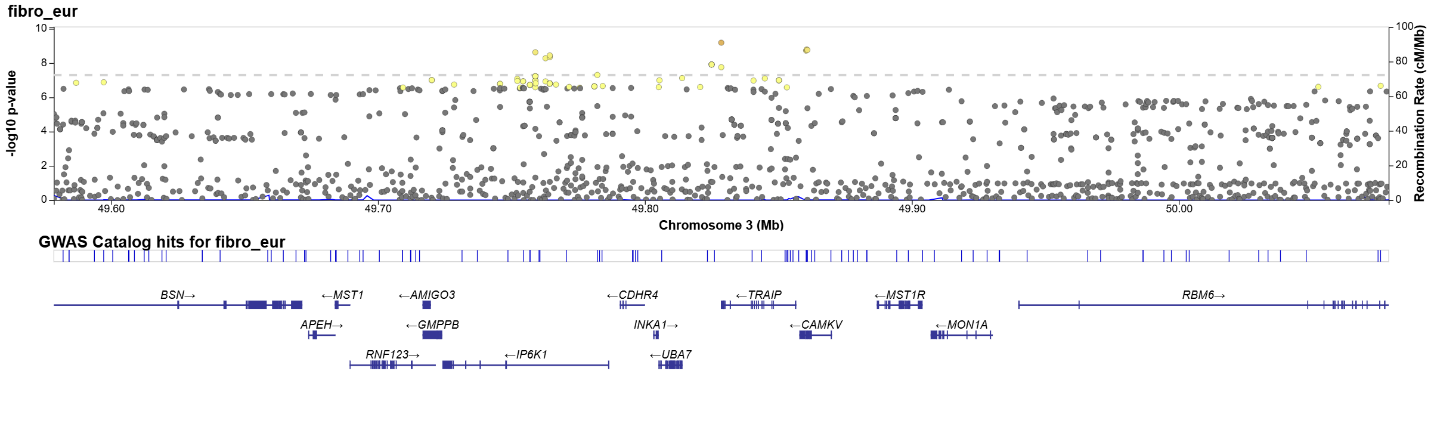
**Figure S11**: regional Manhattan plot: rs71080556 (EUR meta-analysis).

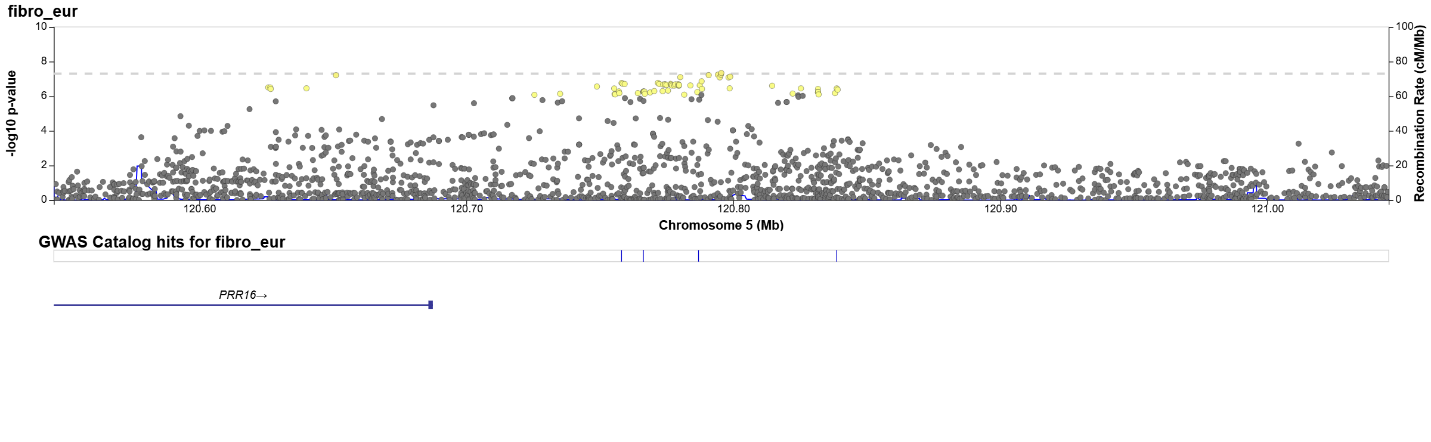
**Figure S12**: regional Manhattan plot: rs56405820 (EUR meta-analysis).

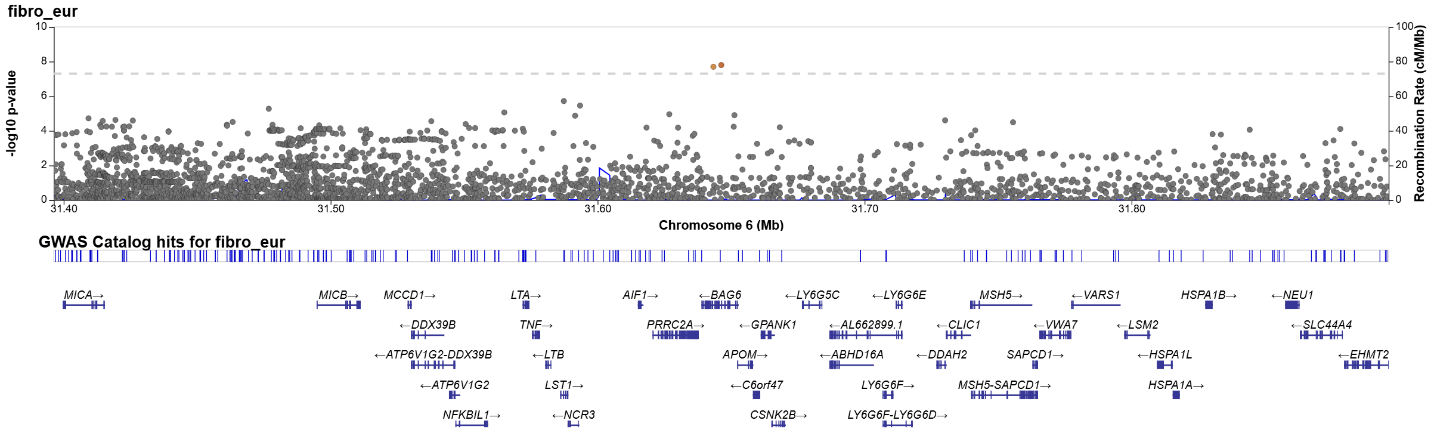
**Figure S13**: regional Manhattan plot: rs2242656 (EUR meta-analysis).

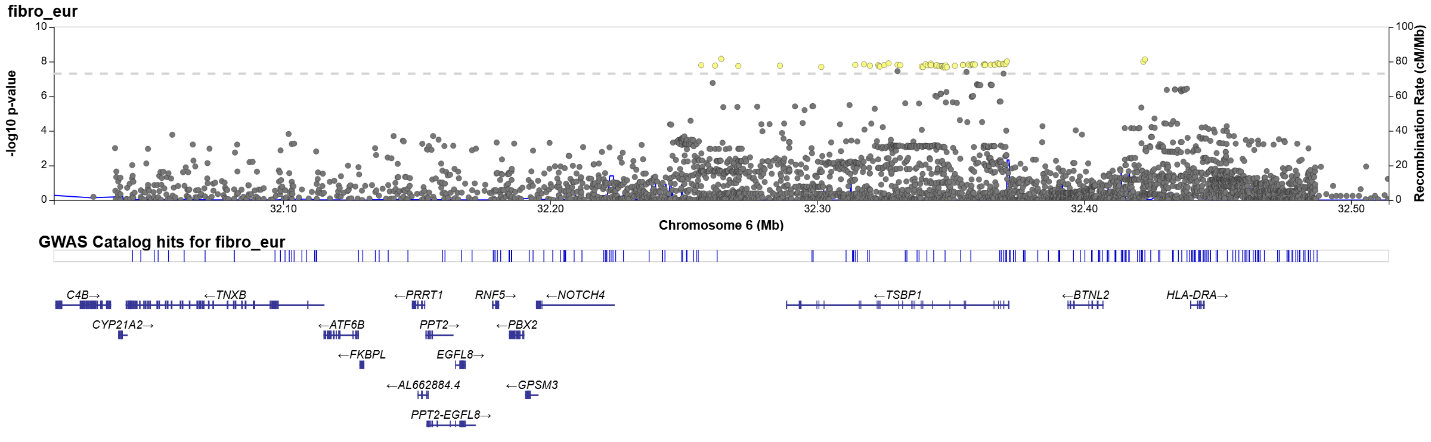
**Figure S14**: regional Manhattan plot: rs9279546 (EUR meta-analysis).

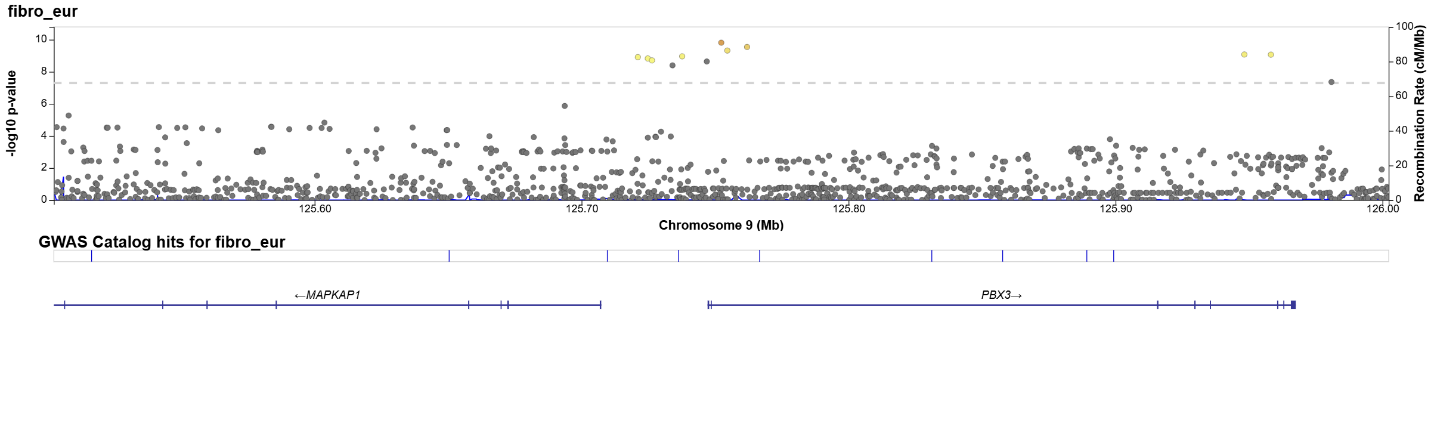
**Figure S15**: regional Manhattan plot: rs6478712 (EUR meta-analysis).

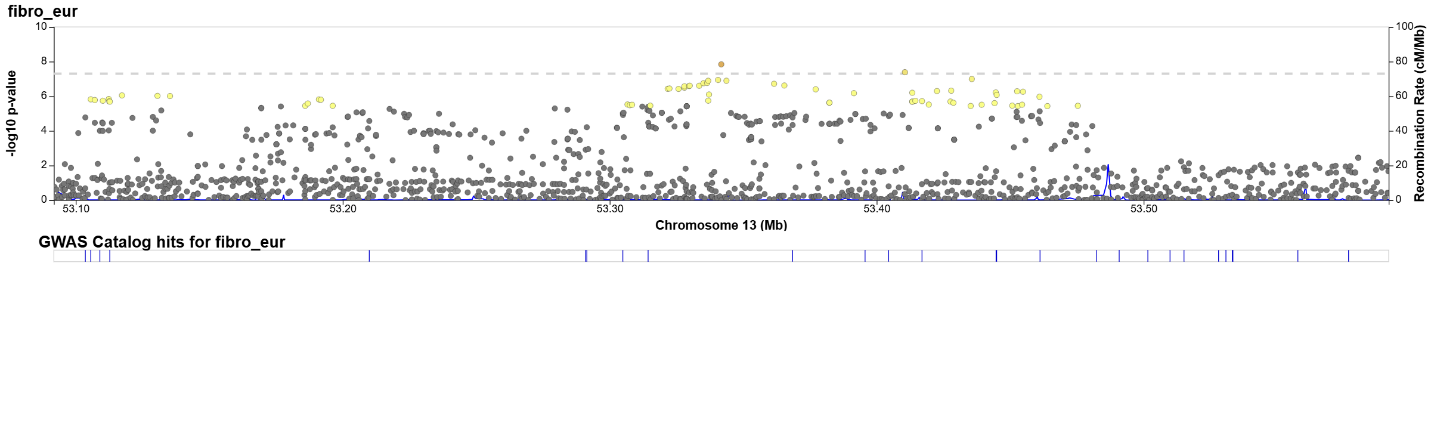
**Figure S16**: regional Manhattan plot: rs2587363 (EUR meta-analysis).

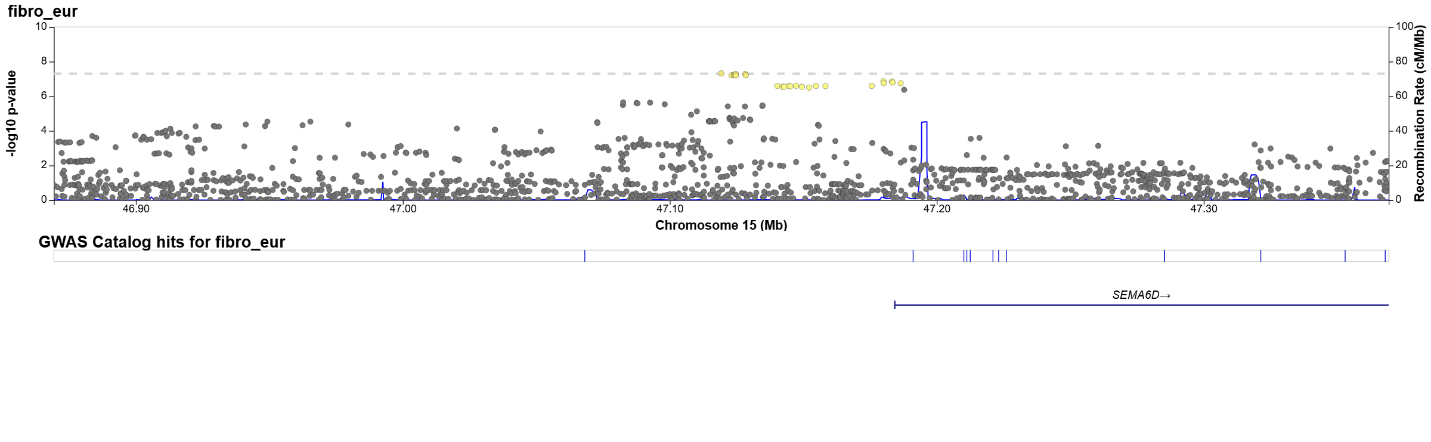
**Figure S17**: regional Manhattan plot: rs181388182 (EUR meta-analysis).

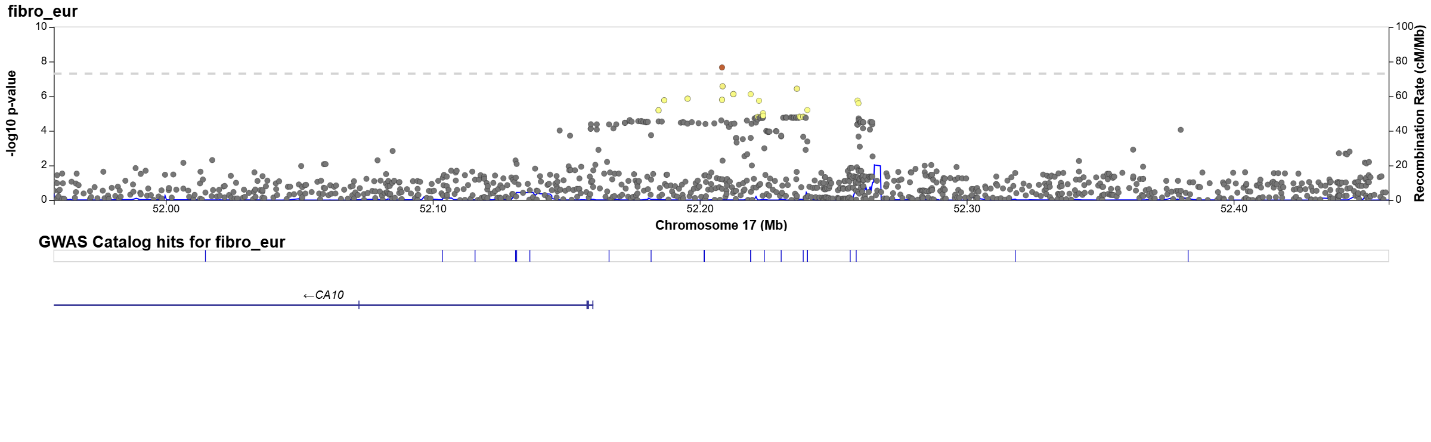
**Figure S18**: regional Manhattan plot: rs11395028 (EUR meta-analysis).

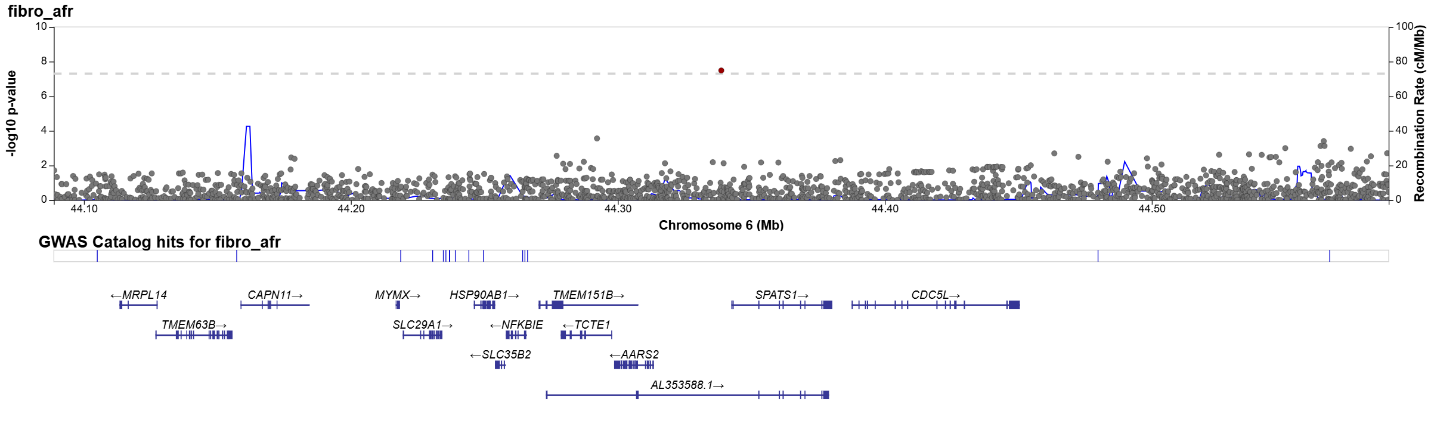
**Figure S19**: regional Manhattan plot: rs186798404 (AFR meta-analysis).

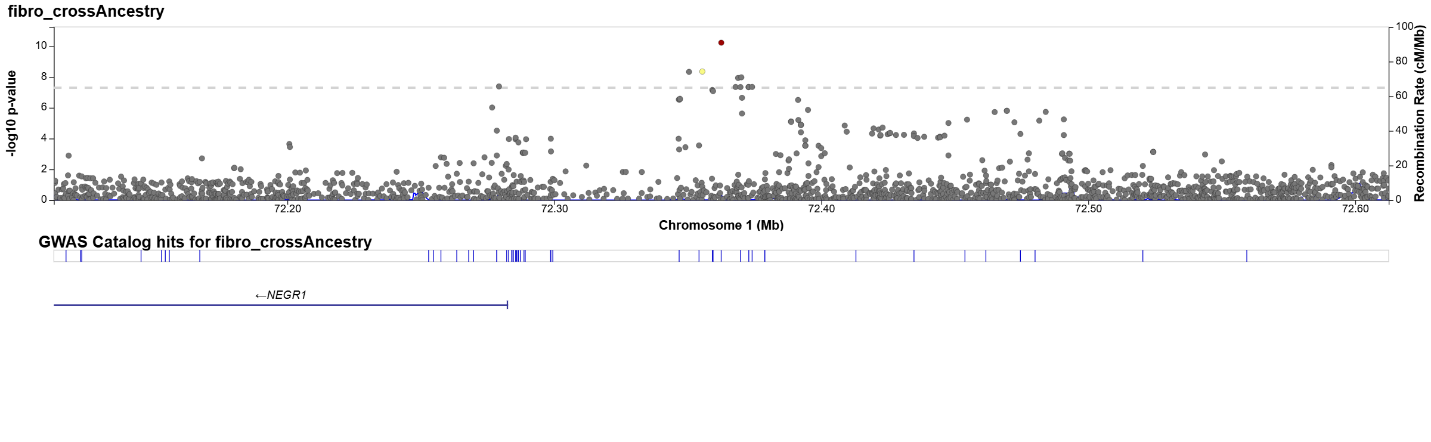
**Figure S20**: regional Manhattan plot: rs10889947 (cross-ancestry meta-analysis).

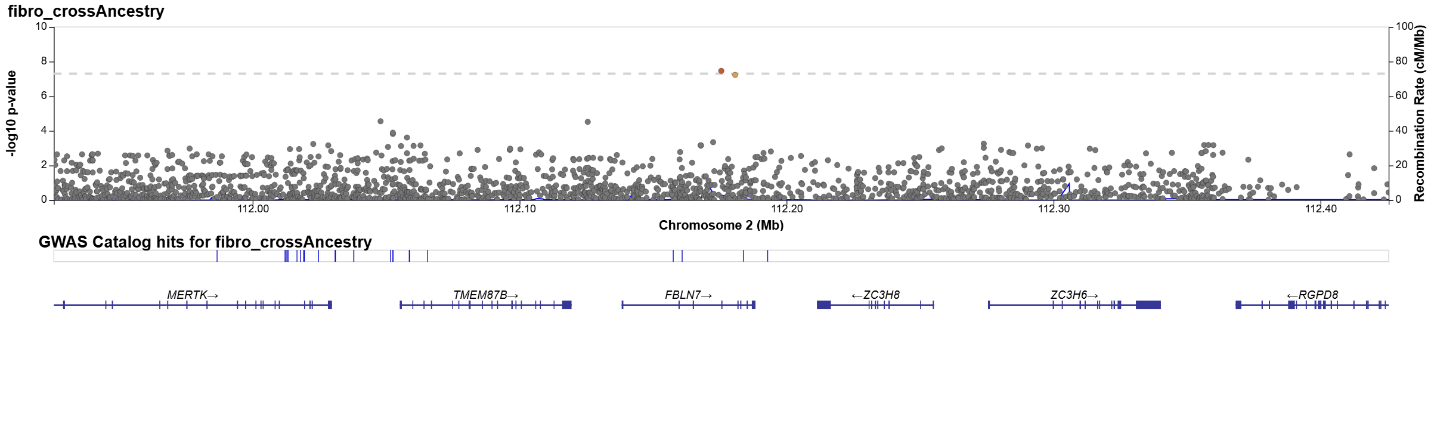
**Figure S21**: regional Manhattan plot: rs72831629 (cross-ancestry meta-analysis).

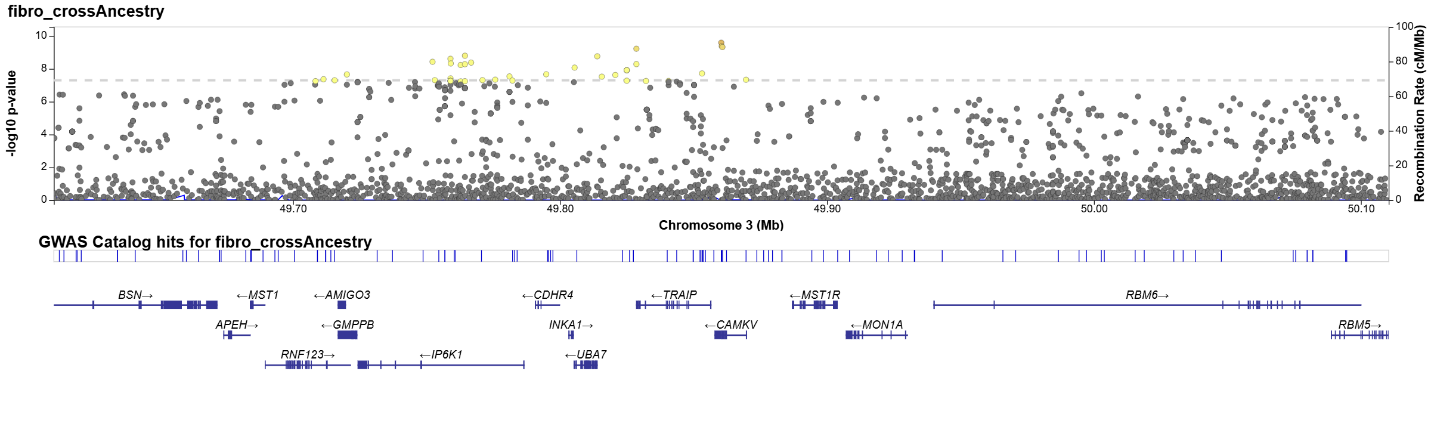
**Figure S22**: regional Manhattan plot: rs2681780 (cross-ancestry meta-analysis).

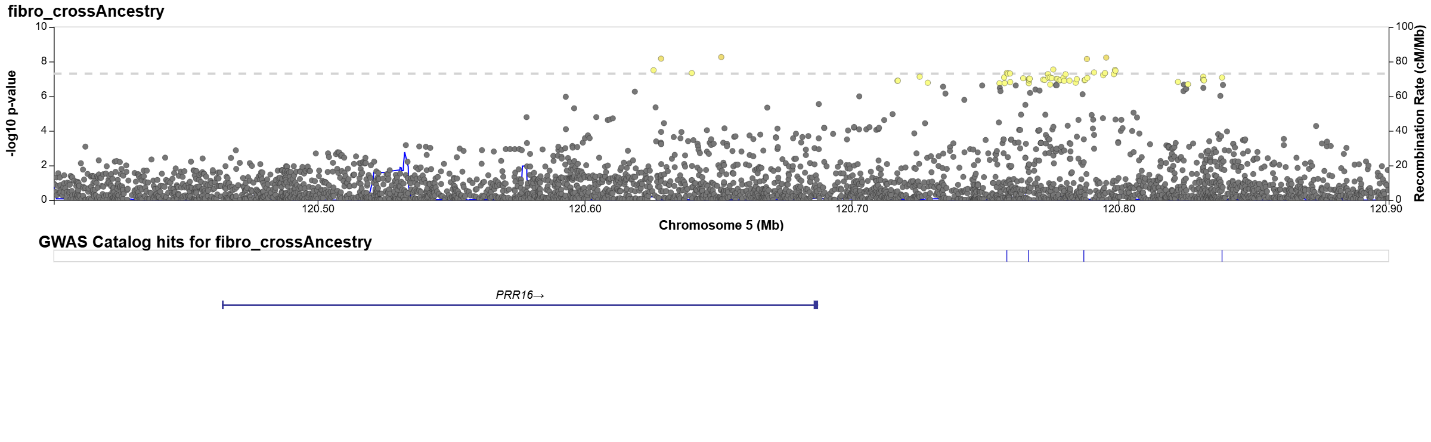
**Figure S23**: regional Manhattan plot: rs190161089 (cross-ancestry meta-analysis).

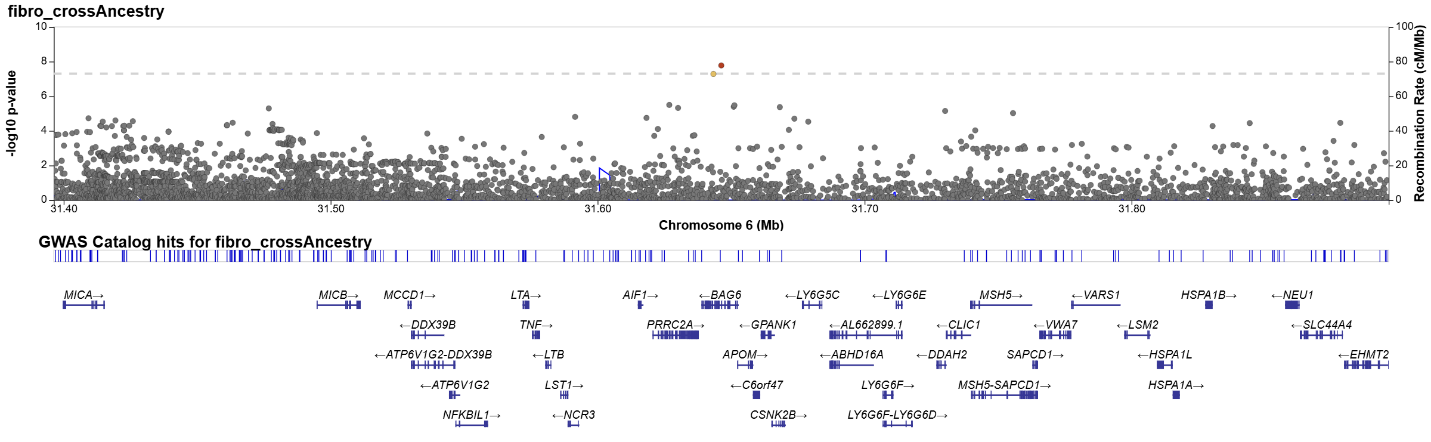
**Figure S24**: regional Manhattan plot: rs2242656 (cross-ancestry meta-analysis).

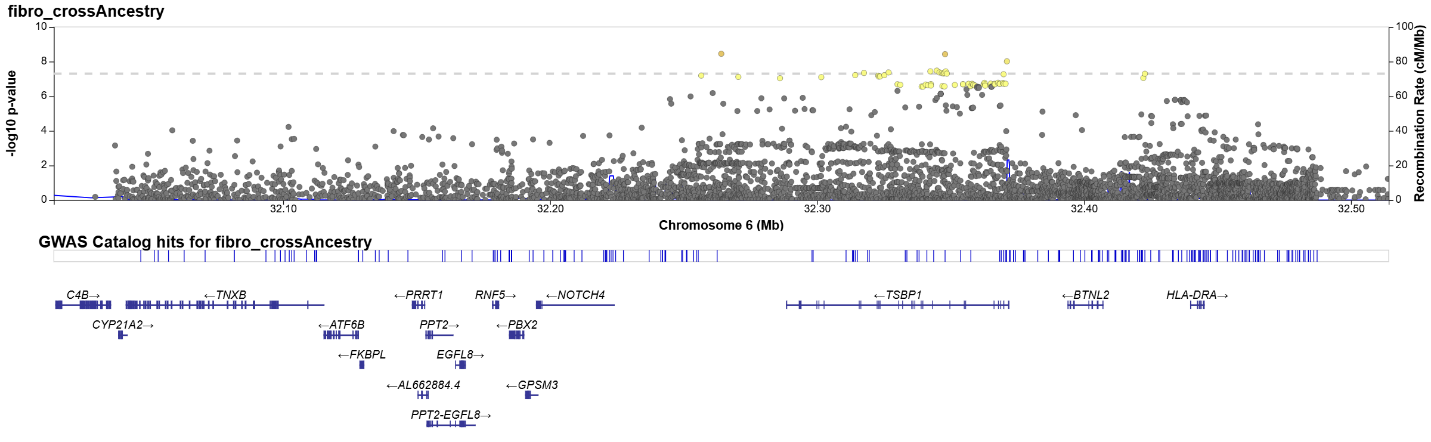
**Figure S25**: regional Manhattan plot: rs9279546 (cross-ancestry meta-analysis).

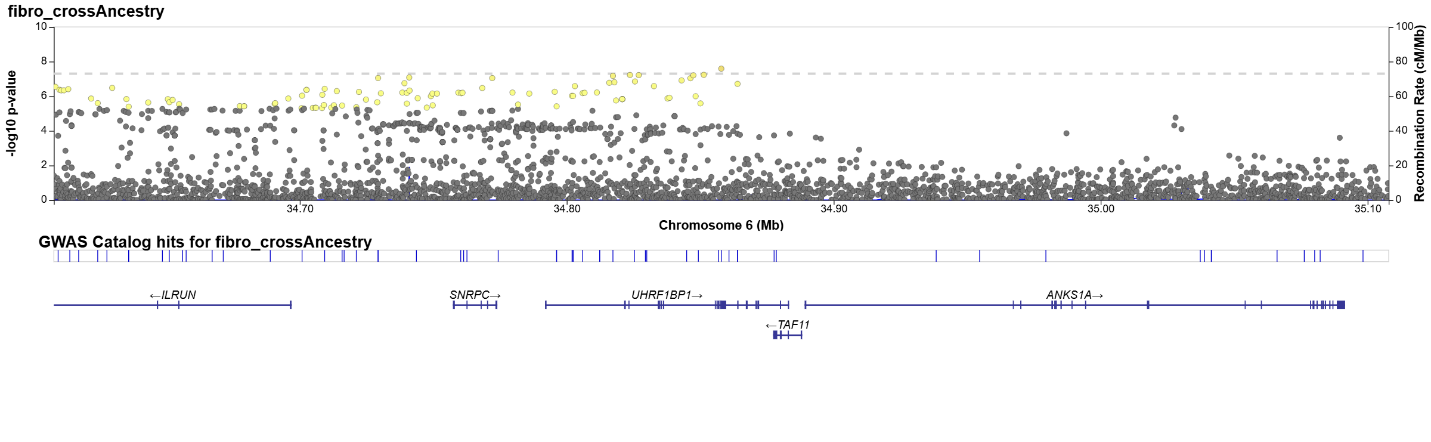
**Figure S26**: regional Manhattan plot: rs16894959 (cross-ancestry meta-analysis).

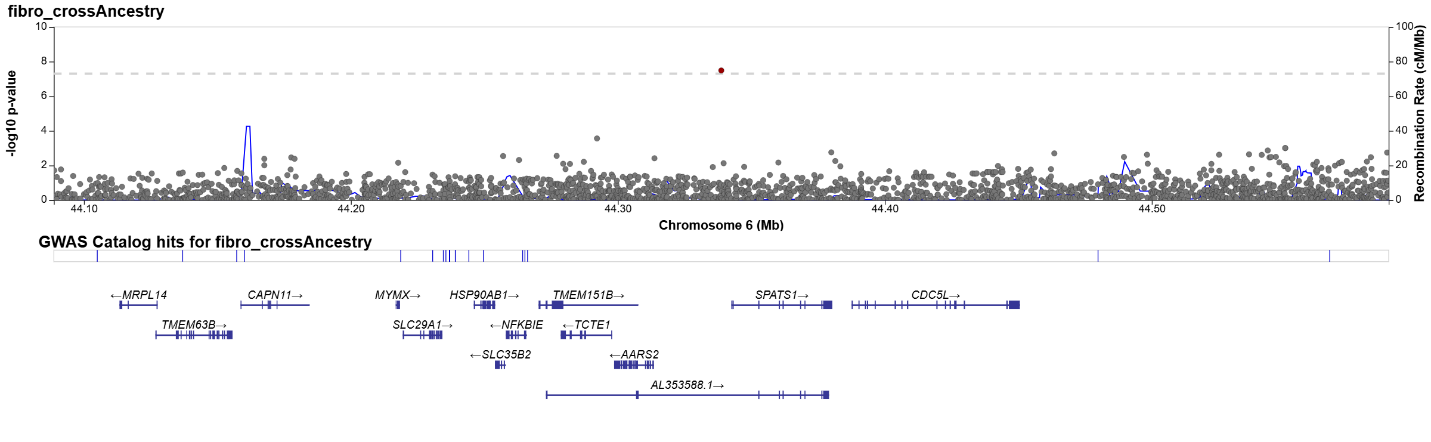
**Figure S27**: regional Manhattan plot: rs186798404 (cross-ancestry meta-analysis).

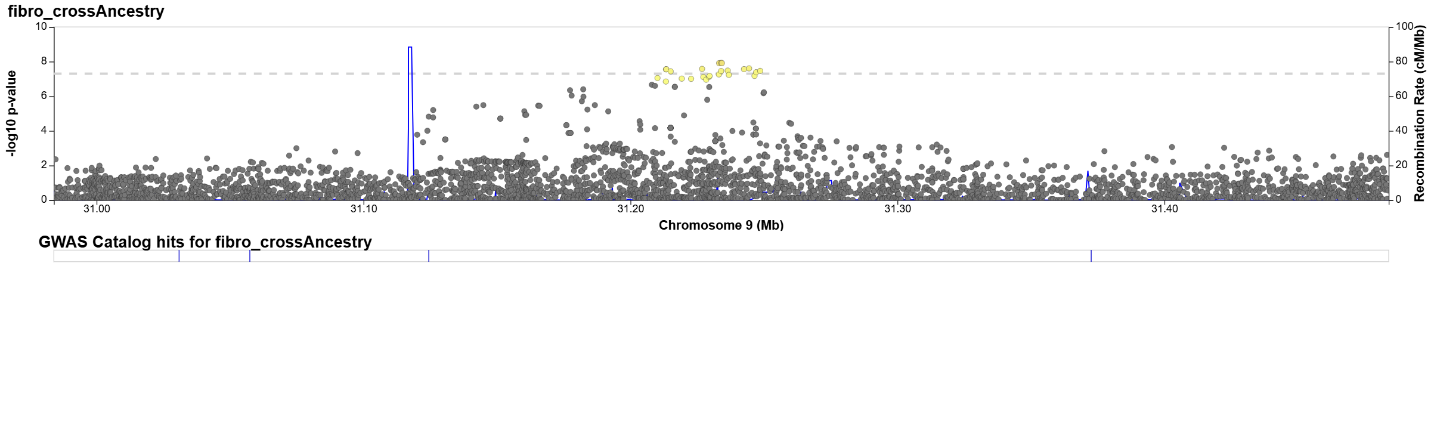
**Figure S28**: regional Manhattan plot: rs12555516 (cross-ancestry meta-analysis).

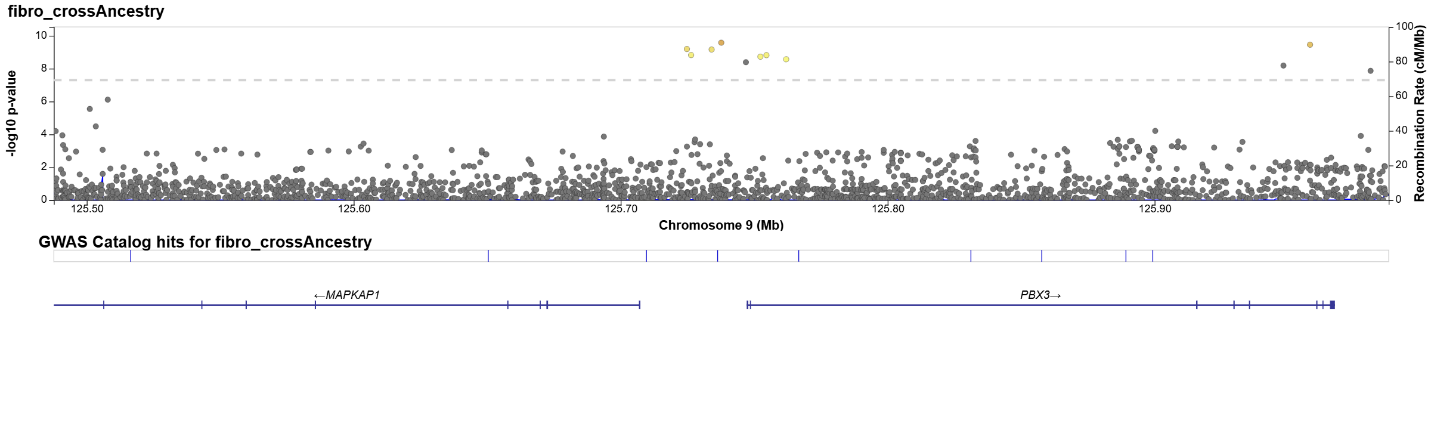
**Figure S29**: regional Manhattan plot: rs10819064 (cross-ancestry meta-analysis).

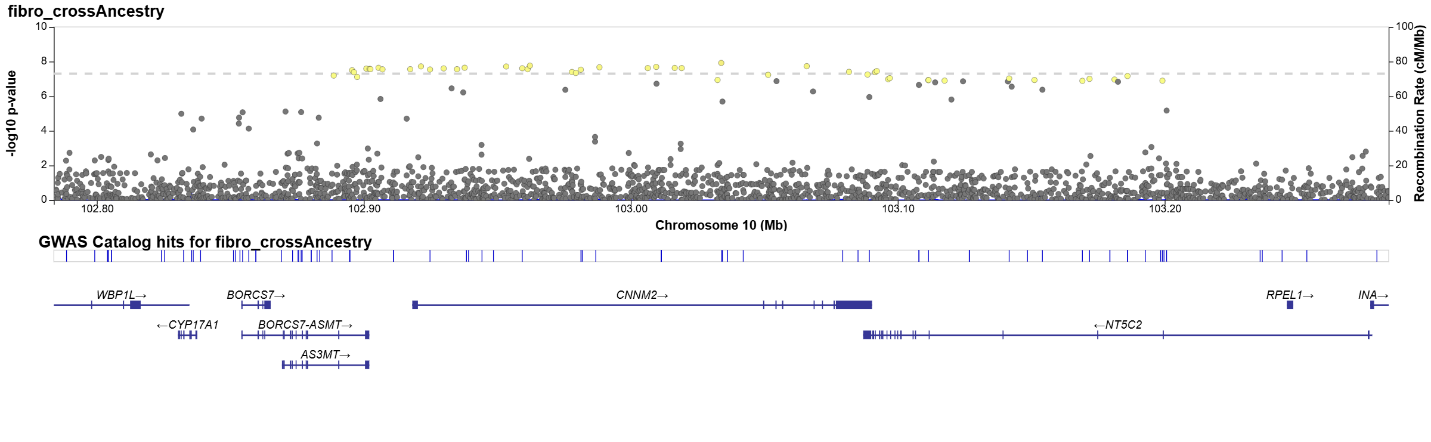
**Figure S30**: regional Manhattan plot: rs75970938 (cross-ancestry meta-analysis).

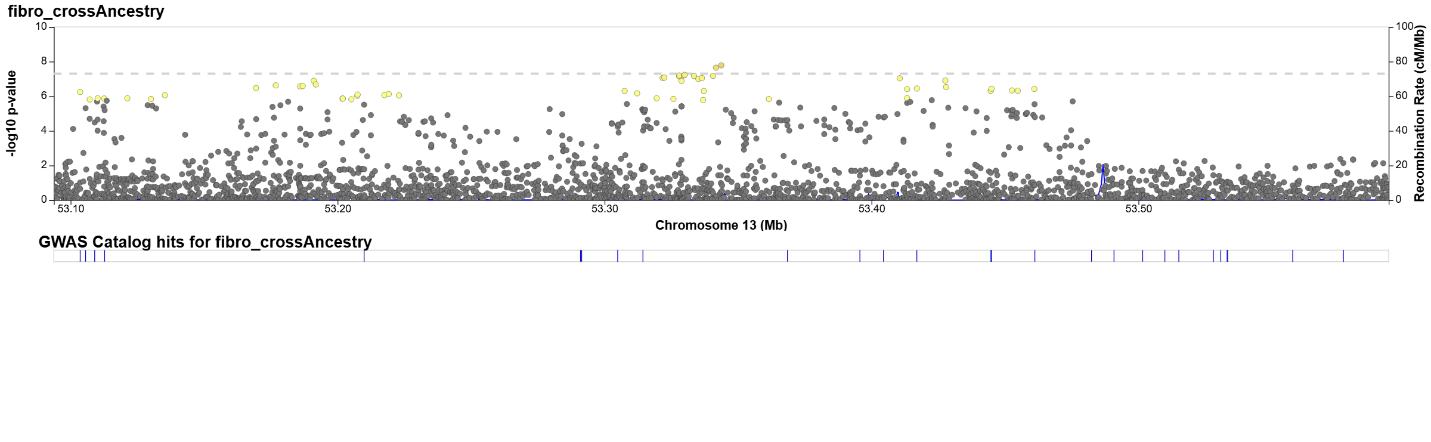
**Figure S31**: regional Manhattan plot: rs9536401 (cross-ancestry meta-analysis).

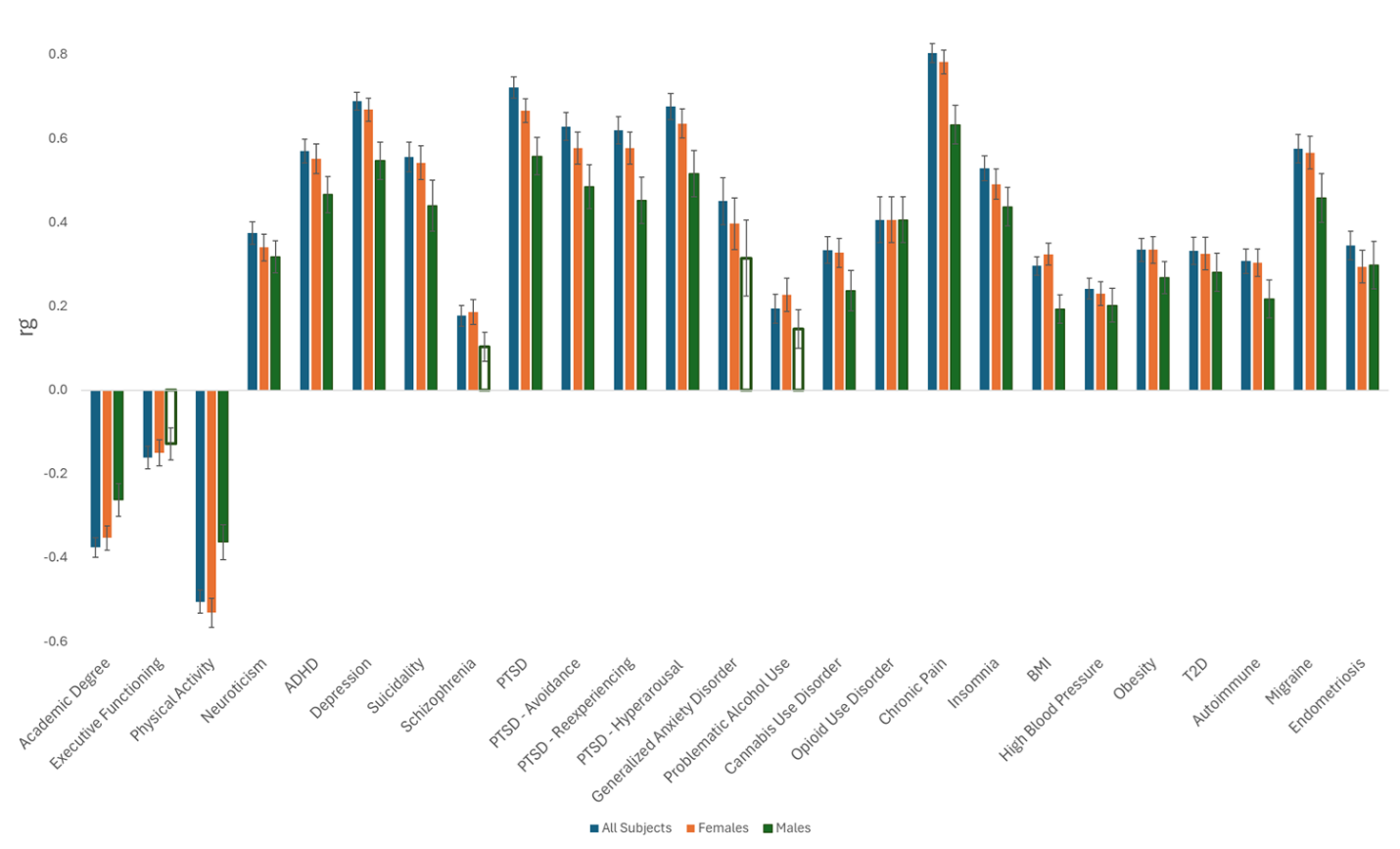

**Figure S32**: Genetic correlations between fibromyalgia and 25 traits of interest in EUR – a comparison between sex-stratified analyses and the main analysis (for the Both Sexes analysis, the data is the same as presented in Figure 2 in the main manuscript). Empty columns represent a non-significant effect [AcaDeg: academic degree; ExecFunc: executive functioning; PhysAct: physical activity; ADHD: attention-deficit/hyperactivity disorder; PTSD: post-traumatic stress disorder; GAD: generalized anxiety disorder; PAU: problematic alcohol use; CanUD: cannabis use disorder; OUD: opioid use disorder; BMI: body mass index; BP: blood pressure; T2D: type 2 diabetes].

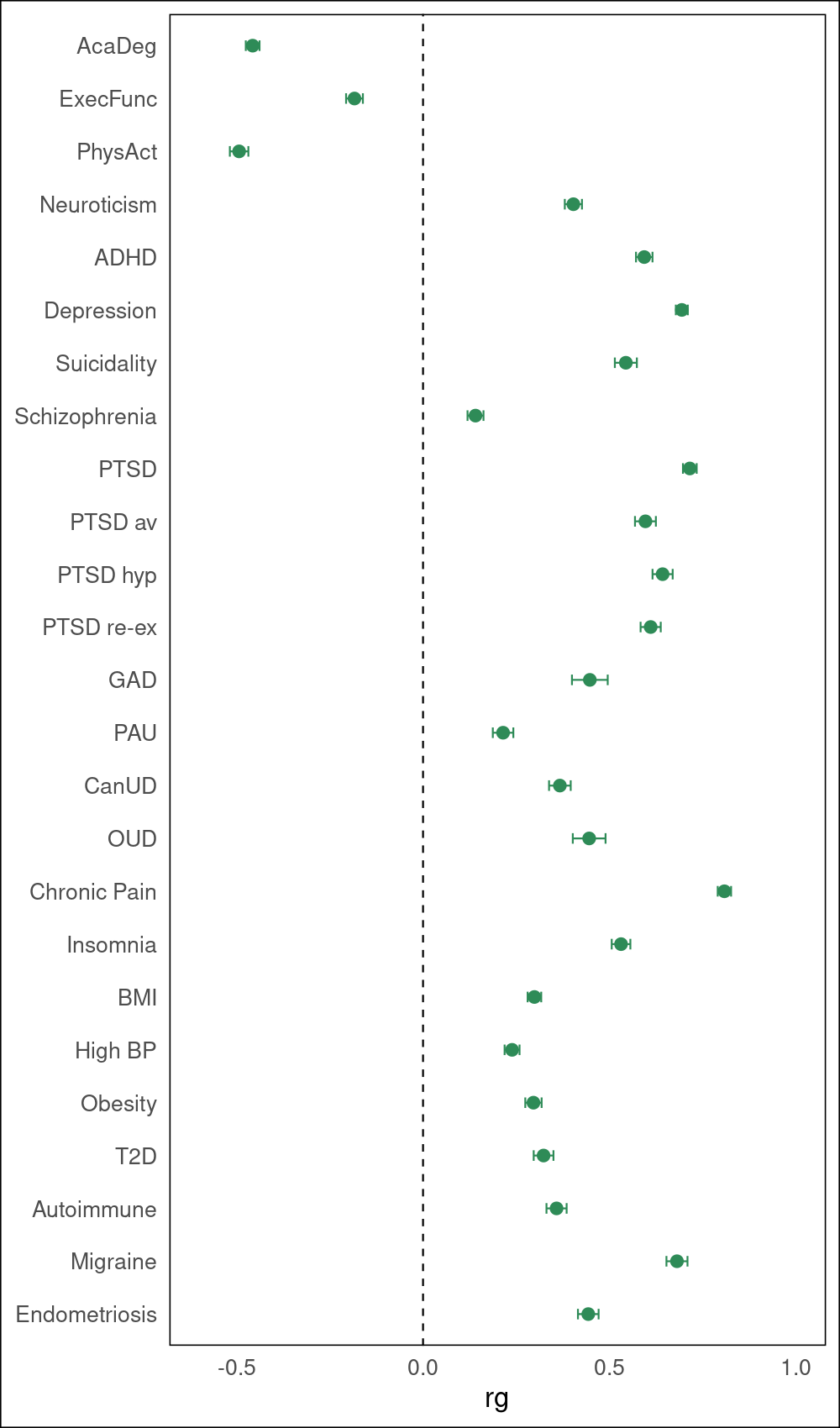

**Figure S33:** Genetic correlations between fibromyalgia MTAG and 25 traits of interest in EUR [AcaDeg: academic degree; ExecFunc: executive functioning; PhysAct: physical activity; ADHD: attention-deficit/hyperactivity disorder; PTSD: post-traumatic stress disorder; av: avoidance; hyp: hyperarousal; re-ex: re-experiencing; GAD: generalized anxiety disorder; PAU: problematic alcohol use; CanUD: cannabis use disorder; OUD: opioid use disorder; BMI: body mass index; BP: blood pressure; T2D: type 2 diabetes].

**Supplementary acknowledgements**

VA Million Veteran Program core acknowledgement
VA Million Veteran Program:
Core Acknowledgements for Publications
May 2024

**MVP Program Office**

- Sumitra Muralidhar, Ph.D., Program Director
US Department of Veterans Affairs, 810 Vermont Avenue NW, Washington, DC 20420

- Jennifer Moser, Ph.D., Associate Director, Scientific Programs
US Department of Veterans Affairs, 810 Vermont Avenue NW, Washington, DC 20420

- Jennifer E. Deen, B.S., Associate Director, Cohort & Public Relations
US Department of Veterans Affairs, 810 Vermont Avenue NW, Washington, DC 20420

**MVP Executive Committee**

- Co-Chair: Philip S. Tsao, Ph.D.
VA Palo Alto Health Care System, 3801 Miranda Avenue, Palo Alto, CA 94304

- Co-Chair: Sumitra Muralidhar, Ph.D.
US Department of Veterans Affairs, 810 Vermont Avenue NW, Washington, DC 20420

- J. Michael Gaziano, M.D., M.P.H. VA Boston Healthcare System, 150 S. Huntington Avenue, Boston, MA 02130

- Elizabeth Hauser, Ph.D. Durham VA Medical Center, 508 Fulton Street, Durham, NC 27705

- Amy Kilbourne, Ph.D., M.P.H.
VA HSR&D, 2215 Fuller Road, Ann Arbor, MI 48105

- Michael Matheny, M.D., M.S., M.P.H.
VA Tennessee Valley Healthcare System, 1310 24th Ave. South, Nashville, TN 37212

- Dave Oslin, M.D.
Philadelphia VA Medical Center, 3900 Woodland Avenue, Philadelphia, PA 19104

- Deepak Voora, MD.
Durham VA Medical Center, 508 Fulton Street, Durham, NC 27705

**MVP Co-Principal Investigators**

- J. Michael Gaziano, M.D., M.P.H.
VA Boston Healthcare System, 150 S. Huntington Avenue, Boston, MA 02130

- Philip S. Tsao, Ph.D.
VA Palo Alto Health Care System, 3801 Miranda Avenue, Palo Alto, CA 94304

**MVP Core Operations**

- Jessica V. Brewer, M.P.H., Director, MVP Cohort Operations
VA Boston Healthcare System, 150 S. Huntington Avenue, Boston, MA 02130

- Mary T. Brophy M.D., M.P.H., Director, VA Central Biorepository
VA Boston Healthcare System, 150 S. Huntington Avenue, Boston, MA 02130

- Kelly Cho, M.P.H, Ph.D., Director, MVP Phenomics
VA Boston Healthcare System, 150 S. Huntington Avenue, Boston, MA 02130

- Lori Churby, B.S., Director, MVP Regulatory Affairs
VA Palo Alto Health Care System, 3801 Miranda Avenue, Palo Alto, CA 94304

- Scott L. DuVall, Ph.D., Director, VA Informatics and Computing Infrastructure (VINCI)
VA Salt Lake City Health Care System, 500 Foothill Drive, Salt Lake City, UT 84148

- Saiju Pyarajan Ph.D., Director, Data and Computational Sciences
VA Boston Healthcare System, 150 S. Huntington Avenue, Boston, MA 02130

- Robert Ringer, Pharm.D., Director, VA Albuquerque Central Biorepository
New Mexico VA Health Care System, 1501 San Pedro Drive SE, Albuquerque, NM 87108

- Luis E. Selva, Ph.D., Director, MVP Biorepository Coordination
VA Boston Healthcare System, 150 S. Huntington Avenue, Boston, MA 02130

- Shahpoor (Alex) Shayan, M.S., Director, MVP PRE Informatics
VA Boston Healthcare System, 150 S. Huntington Avenue, Boston, MA 02130

- Brady Stephens, M.S., Principal Investigator, MVP Information Center Canandaigua
VA Medical Center, 400 Fort Hill Avenue, Canandaigua, NY 14424

- Stacey B. Whitbourne, Ph.D., Director, MVP Cohort Development and Management
VA Boston Healthcare System, 150 S. Huntington Avenue, Boston, MA 02130

**MVP Publications and Presentations Committee**

- Co-Chair: Themistocles L. Assimes, M.D., Ph. D
VA Palo Alto Health Care System, 3801 Miranda Avenue, Palo Alto, CA 94304

- Co-Chair: Adriana Hung, M.D.; M.P.H
VA Tennessee Valley Healthcare System, 1310 24th Ave. South, Nashville, TN 37212

- Co-Chair: Henry Kranzler, M.D. Philadelphia
VA Medical Center, 3900 Woodland Avenue, Philadelphia, PA 19104
